# Reemergence of pathogenic, autoantibody-producing B cell clones in myasthenia gravis following B cell depletion therapy

**DOI:** 10.1101/2022.07.21.22277677

**Authors:** Miriam L. Fichtner, Kenneth B. Hoehn, Easton E. Ford, Marina Mane-Damas, Sangwook Oh, Patrick Waters, Aimee S. Payne, Melissa L. Smith, Corey T. Watson, Mario Losen, Pilar Martinez-Martinez, Richard J. Nowak, Steven H. Kleinstein, Kevin C. O’Connor

## Abstract

Myasthenia gravis (MG) is an autoantibody-mediated autoimmune disorder of the neuromuscular junction. In a subset of patients with MG, muscle-specific tyrosine kinase (MuSK) autoantibodies are present. MuSK MG patients respond well to CD20-mediated B cell depletion therapy (BCDT); most achieve complete stable remission. However, relapse often occurs. To further understand the immunomechanisms underlying relapse, we sought to study autoantibody-producing B cells over the course of BCDT. We developed a fluorescently labeled antigen to enrich for MuSK-specific B cells, which was validated with a novel Nalm6 cell line engineered to express a human MuSK-specific B cell receptor. B cells (≅ 2.6 million) from 12 different samples collected from nine MuSK MG patients were screened for MuSK specificity. We successfully isolated two MuSK-specific IgG4 subclass-expressing plasmablasts from two of these patients, who were experiencing a relapse after a BCDT-induced remission. Human recombinant MuSK mAbs were then generated to validate binding specificity and characterize their molecular properties. Both mAbs were strong MuSK binders, they recognized the Ig1-like domain of MuSK, and showed pathogenic capacity when tested in an AChR clustering assay. The presence of persistent clonal relatives of these MuSK-specific B cell clones was investigated through B cell receptor repertoire tracing of 63,977 unique clones derived from longitudinal samples collected from these two patients. Clonal variants were detected at multiple timepoints spanning more than five years and reemerged after BCDT-mediated remission, predating disease relapse by several months. These results indicate that a reservoir of rare pathogenic MuSK autoantibody-expressing B cell clones survive BCDT and reemerge into circulation prior to manifestation of clinical relapse. This study provides both a mechanistic understanding of MuSK MG relapse and a valuable candidate biomarker for relapse prediction.

## Introduction

Autoimmune myasthenia gravis (MG) is a prototypical autoantibody-mediated disease. Pathogenic autoantibodies in MG target proteins within the neuromuscular junction (NMJ), which interrupts neuromuscular transmission [16, 75]. The largest subset of MG patients have autoantibodies that target the acetylcholine receptor (AChR), but a smaller subset of patients harbor autoantibodies that bind to muscle-specific tyrosine kinase (MuSK) [26], low-density lipoprotein receptor-related protein 4 (LRP4) [25, 51, 84] or agrin [84].

The MG subtypes that are defined by the autoantibody specificity appear clinically similar, but the underlying immunopathology is remarkably distinct [13]. This is well highlighted by comparing the AChR and MuSK subtypes. While AChR MG is driven by the IgG1 and IgG3 subclass [57], MuSK MG is largely governed by the IgG4 subclass that mediates pathology by inhibiting the interaction between MuSK and LRP4 [30], which is essential for MuSK phosphorylation and subsequent effective AChR clustering and signaling. The phenotype of the B cells that produce pathogenic autoantibodies in AChR and MuSK MG also appear divergent. It is currently thought that short-lived plasmablasts are key autoantibody producers in MuSK MG [64, 65], while plasma cells mostly account for the production of AChR autoantibodies [60, 78, 82]. These differences are highlighted by the outcomes reached through the use of biological therapeutics in clinical trials. Specifically, this is evident by the poor response to CD20-mediated (rituximab) B cell depletion therapy (BCDT) in AChR MG [47], in contrast to the significant response in MuSK MG [10, 33]. In clinical practice, most MuSK MG patients achieve complete stable remission following BCDT [10, 44, 48, 49], which often includes reduction of autoantibody titer to undetectable levels. This fits well with an emerging pattern of BCDT efficacy in patients with other diseases mediated by pathogenic IgG4 autoantibodies, such as pemphigus with autoantibodies directed against the desmoglein adhesion molecules [32] and chronic inflammatory demyelinating polyneuropathy with paranodal protein-specific autoantibodies [53]. Notwithstanding the clinically proven efficacy of BCDT, after an initial remission some MuSK MG patients experience relapse, [4, 22, 48, 56], which can associate with an increased frequency of plasmablasts and memory B cells, including populations that express MuSK-specific autoantibodies [65, 68]. Not all of these B cells develop *de novo* after BCDT, as a proportion of B cell clones persist through the treatment [31]. However, it is not clear how pathogenic MuSK autoantibody-expressing B cell clones behave during a clinical course, as it includes phases of relapse and periods of remission induced by repeated BCDT treatment cycles.

To that end, we sought to isolate MuSK-specific autoantibody-producing B cells, validate their specificity, and determine whether these B cells are present in BCDT-treated patients over a period of time. We developed a monomeric fluorescently labeled MuSK antigen to enrich for MuSK-specific B cells and authenticated this antigen bait using a novel B cell line (Nalm6 cells) that expressed a human MuSK-specific B cell receptor (BCR). We found that MuSK-specific B cells are exceptionally rare in the circulation, yet we were able to isolate two distinct clones from two different patients, the specificity of which was validated through testing of human recombinant monoclonal autoantibodies (mAbs). We collected upwards of 149,000 B cell receptor sequences from serial samples of these two patients to search for clonal variants. We found that MuSK-specific B cell clones persisted through repeated rounds of BCDT and reemerged prior to clinical relapse in association with increased autoantibody titers.

## Material and methods

### Human specimens

This study was approved by the Human Investigation Committee at the Yale School of Medicine (clinicaltrials.gov || NCT03792659). Informed written consent was obtained from all patients. All MuSK MG patients met definitive diagnostic criteria for MG, including positive serology for MuSK autoantibodies.

### Fluorescently labeled MuSK, Nalm6 cells, and cell sorting

Recombinant human MuSK and the negative control protein recombinant human growth hormone (hGH) (BioLegend; 778006) were fluorescently labeled using the Alexa Fluor™ 647 Microscale Protein Labeling Kit (Invitrogen; A30009). Nalm6 cells (CRL-3273™) and Nalm6 cells containing the MuSK-specific MuSK3-28 B cell receptor (Nalm6_3-28) were cultured in RPMI 1640 media containing 10% FBS, 1% P/S and 1% HEPES. On the day of the CBA for the validation of the MuSK reagent, the fluorescently labeled MuSK was added at a final concentration of 10, 1, 0.1 or 0.01 μg/ml. For sorting MuSK-reactive B cells, B cells were enriched from cryopreserved PBMCs using negative selection beads (Stemcell Technologies; 19554). They were incubated with live/dead stain, then stained for 30 minutes on ice with 1 μg/ml of fluorescently labeled MuSK antigen together with fluorescently labeled antibodies against CD3 (BD Biosciences, V500; UCTH1), CD14 (Invitrogen, Pacific orange; TUK4), CD19 (BioLegend, PE Cy7; SJ25C1), CD27 (BD Biosciences, PE; M-T271), IgD (BD Biosciences, FITC; IA6-2), IgM (BioLegend, PerCP/Cyanine5.5; MHM-88) and CD38 (BioLegend, BV421; HB-7) using manufacturer’s recommended dilutions. The cells were sorted on an FACSAria (BD Biosciences) instrument. For general B cell immunophenotyping, B cells were defined as live CD3–CD14–CD19+ cells, memory B cells as live CD3–CD14–CD19+CD27+CD38– cells, and antibody-secreting cells (plasmablast phenotype) as CD3–CD14–CD19+CD27+CD38hi. We sorted for CD3-CD14-CD19+IgD-CD27+IgM-MuSK+ cells for the single cell B cell culture experiments.

### B cell culture, molecular cloning, and IgG subclass determination

MS40L^lo^ cells (kindly provided by Drs. Garnett Kelsoe and Dongmei Liao of Duke University; [40]) were maintained in IMDM media (Invitrogen, 12440-053) supplemented with 10% FCS (Thermo Scientific, SH30070.03), 1% Pen/Strep (Invitrogen, 15140-122) and 55 µM 2-ME (Invitrogen, 21985). For B cell culture the cells were suspended in B cell media (RPMI, 10% FBS, 1% Pen/Strep, 1% HEPES, 1% Sodium pyruvate, 1% MEM NEAA and 55 µM of 2-ME) and plated at a concentration of 3 x 10^3^ cells per well into 96 well plates. The B cell media was changed the following day to contain additionally 50 ng/ml IL-2 (Peprotech 100-02), 10 ng/ml IL-4 (Peprotech 200-04),10 ng/ml IL-21 (Peprotech 200-21) and 10 ng/ml BAFF (Peprotech 310-13) and single B cells were sorted into each well. The culture was maintained for 20-30 days, then the supernatant was harvested and screened for MuSK-reactivity using a MuSK-specific cell-based assay (CBA). The RNA of cells in MuSK antibody positive wells was purified using the RNeasy Plus Mini Kit (Qiagen), followed by cDNA synthesis, single-cell PCR, IgG subclass determination and molecular cloning into the corresponding heavy, kappa or lambda vectors as previously described [68, 76]}.

### Recombinant expression of MuSK and human mAbs and Fabs

The extracellular domain of MuSK (AA 22-452) was produced in stably transfected Drosophila S2 cells kindly provided by Dr. Patrick Waters (University of Oxford) as previously described [68]. The 2E6 and 6C6 mAbs and were expressed as IgG1 subclass whole antibodies as previously described [68]. The negative control mAb D12 was generated from patient MuSK MG-3. It was derived from single cell sorting and subsequent single cell PCR. The sort enriched for IgG4-specific memory B cells/plasmablasts (CD3^neg^, CD14^neg^, CD19^+^, IgD^neg^, CD27^+^, IgG4^+^ (biotinylated IgG4-specific clone MH164-1, with APC streptavidin (Biolegend #405207) used for detection). The antibody subclass usage of this clone was verified as IgG4 through subclass PCR. The D12 mAb was expressed as an IgG4 subclass whole antibody, and then used as a negative control as no binding to MuSK via CBA was observed. The antigen binding fragments (Fabs) of 2E6 and 6C6 were expressed in a human Fab expression vector (see below for heavy chain plasmid description) using the same expression system as described for the mAbs. Protein G Sepharose® 4 Fast Flow beads were used for mAb purification and the 6XHis-tagged Fabs and MuSK were affinity purified using HisPur^TM^ Cobalt Resin according to manufacturer’s protocol.

### Immunofluorescence of mouse muscle sections

The binding of the mAb 2E6 and 6C6 to mouse MuSK was performed as previously described [68]. Briefly, positive control AChR-specific mAb 637 [18], 6C6 and 2E6 mAbs were added at a concentration of 4 μg/mL each combined with Alexa Fluor 647–conjugated α-bungarotoxin (1:300, B35450, Thermo Fischer Scientific). Human Fc-γ–specific Alexa Fluor 488–conjugated goat F(ab′)_2_ (3 μg/mL, 109-546-170, Jackson ImmunoResearch) was subsequently added. Endplates were identified using the immunofluorescence of the α-bungarotoxin staining. Triple-fluorescent photomicrographs of the endplate regions were acquired using μManager software ver2.0 [70] on an Olympus BX51WI spinning-disk confocal fluorescence microscope with a Hamamatsu EM-CCD C9100 digital camera. Endplates were analyzed using ImageJ software (NIH) as described [17, 70].

### Autoantibody binding cell-based assay

HEK293T cells were transfected with either full-length MuSK-GFP (kindly provided by Drs. David Beeson, Angela Vincent and Patrick Waters, Neurosciences Group at the Weatherall Institute of Molecular Medicine, University of Oxford) or different ectodomain variants of MuSK-GFP (previously described in [68]). The cell-based assay (CBA) was performed as previously decribed [14]. The autoantibody titer of human sera was measured using 10 two-fold dilutions ranging from 1:20 to 1:10240. The binding of each mAb was detected with Alexa Fluor^®^-conjugated AffiniPure Rabbit Anti-Human IgG, Fcγ (309-605-008, Jackson Immunoresearch) on a BD LSRFortessa^®^ (BD Biosciences). FlowJo software (FlowJo, LLC) was used for analysis.

### Human Fab expression vector construction

A human Fab expression vector was engineered from our human IgG1 heavy chain expression vector [50], following a human Fab VH vector design demonstrated to work with mammalian antibody expression [73]. The region coding for the IgG1 constant region was modified to terminate near the junction of the CH1 region and the upper hinge (TKVDKKV - EPKSC). At this region a linker sequence (GS) was added followed by a 6XHis-tag, then a stop codon (TKVDKKV – EPKSC – GS – HHHHHH - stop). A synthetic DNA fragment (gBlock™, Integrated DNA Technologies) coding for the modified constant region was amplified by PCR (GoTaq® DNA Polymerase; Promega (M3001)) and cloned into the original human IgG1 heavy chain expression vector at the Apa I site—located at the beginning of the CH1—and a Bam HI site, which is located at the end of CH3 immediately downstream of the stop codon. The sequence integrity of this new human Fab heavy chain expression vector was confirmed by both Sanger sequencing of the insert and sequencing of the entire plasmid with the Oxford Nanopore platform (Plasmidsaurus). The variable regions coding for the heavy chains of the mAbs 2E6, 6C6, MuSK1A and IgG4 D12 were then subcloned into the Fab expression vector at the Afe I and Apa I sites following standard procedures. The plasmids were transformed into NEB® 5- alpha Competent E. coli (New England BioLabs, Inc.). Plasmid DNA was then isolated with the QIAprep Spin Miniprep Kit (Qiagen) and sequenced by Sanger sequencing to confirm the presence of each specific variable region.

### Polyreactivity, HEp-2 ELISA and AChR clustering assay

Recombinant human mAbs were tested for polyreactivity on microplates coated with 20 µg/ml double-stranded DNA (dsDNA), 10 µg/ml lipopolysaccharide (LPS), or 15 µg/ml recombinant human insulin (all purchased from SIGMA-Aldrich) using a previously described approach [76]. Highly polyreactive antibody ED38 was used as positive control [63]. Purified recombinant IgGs were tested for autoreactivity on a commercially available human epithelial type 2 (HEp-2) cell lysate ELISA kit (INOVA) according to the manufacturer’s instructions with minor modifications that have been described previously [85]. The ELISA plates were read using a PowerWave XS (BIO-TEK). The C2C12 AChR clustering assay was performed as previously reported [14, 68].

### Bulk library preparation

RNA was isolated from frozen PBMCs using the RNeasy Mini Kit (Qiagen, 74104) according to manufacturer’s instructions. BCR libraries were either generated using the NEBNext Immune Sequencing Kit (NEB) as previously published [31] or the SMARTer® Human BCR IgG IgM H/K/L Profiling Kit (Takara Bio USA, Inc.) using the primers targeting IgA, IgG and IgG-subclasses. Four libraries were pooled in equimolar amounts and prepared for sequencing on the Pacific Biosciences (PacBio) Sequel II machine by B Cell Receptor Repertoire SMRTbell® Library (PacBio) preparation.

### Single-cell library preparation

B cells were enriched from cryopreserved PBMCs using negative selection beads (Stemcell Technologies; 19554). They were incubated with live/dead stain, then stained for 30 minutes on ice with fluorescently labeled antibodies against CD3 (BD Biosciences, V500; UCTH1), CD14 (Invitrogen, Pacific orange; TUK4), CD19 (BioLegend, PE Cy7; SJ25C1), CD27 (BD Biosciences, PE; M-T271), IgD (BD Biosciences, FITC; IA6-2), IgM (BioLegend, PerCP/Cyanine5.5; MHM-88) and CD38 (BioLegend, BV421; HB-7) using manufacturer’s recommended dilutions. The cells were sorted on an FACSAria (BD Biosciences) instrument and the population of CD3-CD14-CD19+IgD-CD27+IgM-CD38+ was collected for subsequent analysis by 10x Genomics. Sorted B cells were then loaded into the Chromium Controller (10x Genomics). Single-cell gene expression libraries were prepared using the Chromium Single-cell 5′ Reagent Kit (10x Genomics; V 2.0) according to manufacturer’s instructions. Samples were sequenced on the NovaSeq 6000 Sequencing System (Illumina) with HiSeq paired-end, 150bp reads for 10x Single cell BCR (BCR libraries) and 10x Single cell 5 Prime (gene expression).

### Single B cell RNA-seq analysis

Single cell RNA-seq gene expression information from all subjects was processed using Seurat v4.1.1 [23] in R v4.1.0. To remove apoptotic or lysing cells, cells with a ≥10% of RNA transcripts from mitochondrial genes were excluded. To exclude poor quality cells, only cells with reads from > 400 features were retained. Read counts were log-normalized using a scaling factor of 10^4^. To account for variability in gene expression, log-normalized read counts were then scaled and centered for each feature. The top 2000 variable genes were then identified using Seurat’s “vst” method. V, D, and J genes from the IGH, IGL, and IGK loci were removed so that the properties of the BCR expressed by the cell would remain independent of the cluster to which it was assigned. Seurat’s IntegrateData function was then used to combine data from both sequencing runs. Integration was performed using the previously identified top variable genes of each run, and the first 20 dimensions. Following integration, variable gene expression values were re-scaled and centered. This data was then reduced to the first 20 principal components. To annotate B cell subtypes, cells were clustered by Seurat’s shared nearest neighbor clustering algorithm with a resolution of 0.5. The B cell subtype of each cluster was then determined by gene expression correlations to cell types in the immunoStates database [5]. This resulted in 3 clusters identified as plasmablasts, 6 identified as memory B cells, and 2 identified as naïve B cells. These cell type annotations were verified using known marker genes for plasmablasts (PRDM1, XBP1), memory B cells (CD24, TNFRSF13B) and naïve B cells (IGHD, IL4R, TCL1A). One cluster of plasmablasts was further identified as “proliferating plasmablasts” due to high expression of MKI67.

### B cell receptor sequence processing

Bulk and single cell B cell receptor sequences were obtained from four different data sources: 10X Genomics single cell RNAseq + BCR sequencing, bulk heavy-chain only BCR from New England Biolabs NEBNext sequencing kits, Takaras SMARTer® Human BCR IgG IgM H/K/L Profiling Kit, and previously published, processed bulk BCR sequences from the same patients, which we previously reported [43]. All BCR repertoire sequence data were analyzed using the Immcantation (www.immcantation.org) framework. Heavy and light chain BCR sequence data from 10X Genomics scRNA-seq + BCR sequencing began with the filtered V(D)J contigs from Cell Ranger. To obtain V and J gene assignments, these contigs were aligned to the IMGT GENE-DB [19] (v3.1.23, obtained August 3, 2019) human germline reference database using IgBlast v1.13.0 [81]. Preprocessing of NEBNext BCR sequences was performed using pRESTO v0.6.2 [72]. Quality control was first performed by removing all reads with a Phred quality score < 20. Reads which did not match to a constant region primer (maximum error rate 0.2) or template switch sequence (maximum error rate 0.5) were discarded. A unique molecular identifier (UMI) was assigned to each read using the first 17 nucleotides following the template switch site. Sequences within each UMI group were then collapsed into consensus sequences. Clusters with error rates exceeding 0.1 or majority isotype found in less than 60% of sequences were discarded. Positions containing more than 50% gap sequences were removed from the consensus. Mate-pairs were assembled into sequences with a minimum overlap of 8 nucleotides and a maximum error of 0.3. Isotypes were then assigned by local alignment of the 3’ end of the assembled sequences to known isotype-constant region sequences with a maximum error rate of 0.3. Duplicate sequences were collapsed except if assigned to different isotypes. Sequences represented by a single reconstructed mate-pair were discarded. To obtain V and J gene assignments, remaining sequences were aligned to the IMGT GENE-DB human reference database (v3.1.23, obtained August 3, 2019) using IgBlast v1.13.0.

### Pacific Biosciences B cell receptor sequence preprocessing

HiFi reads generated from single molecule, real-time (SMRT) sequencing data were first demultiplexed using the Lima tool (https://github.com/PacificBiosciences/pbbioconda) based on the Illumina indices integrated during library construction. Sample-level demultiplexed reads were then further processed using pRESTO [72], following a similar workflow to that used for the NEBNext-generated libraries. Briefly, reads with Phred quality scores < 20 were removed, followed by the identification of the universal primer on the 5’ end of each read; reads with primer annotation error rates exceeding 0.3 were discarded. Sequences with the same UMIs were then clustered and aligned to generate collapsed consensus sequences representing each unique UMI. Consensus reads with < 2 representative sequences from the dataset were removed. The remaining reads for each sample were assigned to respective IGHV, IGHD, and IGHJ genes using IgBLAST with the IMGT database as the reference (downloaded February 21, 2022).

### B cell clonal analysis

BCR sequences from all data sources were pooled together and grouped by subject. Nonproductive heavy chains were removed. Light chain sequences were excluded from clonal clustering analysis. To limit low-coverage sequences, all sequences with fewer than 300 unambiguous nucleotide characters (ATCG) were discarded. Novel IGHV alleles and subject-specific IGHV genotypes were inferred using TIgGER v1.0.0 [15]. To identify B cell clones, sequences were first partitioned based on common V and J gene annotations, as well as junction length. Within these groups, sequences differing from one another by a Hamming distance threshold of 0.15 within the junction region were clustered into clones using single linkage hierarchical clustering [20, 21] implemented in Change-O v1.2.0 [21]. This Hamming distance threshold was determined by manual inspection of distance to the nearest sequence neighbor plot using shazam v1.1.0 [80]. Three clones containing both high-throughput BCR sequences and monoclonal antibody sequences were identified. Unmutated germline V and J gene sequences were then reconstructed for each of these clones using the createGermlines function within dowser v1.0.0 [27]. To infer lineage trees, tree topologies, branch lengths, and substitution model parameters were estimated first under the GY94 model [46] and then under the HLP19 model [28]. All lineage tree analysis used IgPhyML v1.1.4 [28] and dowser v1.0.0 [27]. Trees were visualized using dowser v1.0.0, ggtree v3.0.4 [83], and ggplot v3.3.6 [77]. All B cell clonal analyses were performed using R v4.1.0. Scripts for performing B cell receptor sequence processing and analyses are available at https://bitbucket.org/kleinstein/projects.

### Statistics

Statistical significance was assessed with Prism Software (GraphPad; version 8.0) by multiple comparison ANOVA with Dunnett’s correction for AChR clustering in the C2C12 assay.

### Data Availability

Anonymized data will be shared on request from qualified investigators and completion of materials transfer agreements.

## Results

### Validation of fluorescently labeled MuSK

We generated a fluorescently labeled human MuSK ectodomain to specifically identify and isolate MuSK-specific B cells from patient samples **(Supplement Fig. 1)**. We verified the utility of the labeled MuSK by testing it with a human B cell precursor line (Nalm6), which was modified to express a MuSK-specific human recombinant mAb (MuSK3-28) in the form of a BCR [11, 65, 68]. Fluorescently labeled recombinant human growth hormone (hGH) was used as a negative control antigen. The fluorescently labeled MuSK was tested over a broad range of concentrations (0.01, 0.1, 1.0 and 10 μg/mL), and showed strong binding to the Nalm6 cells, which expressed the MuSK3-28 BCR **(Supplement Fig. 1a, b)**. No binding of either antigen was observed with the unmodified Nalm6 cells **(Supplement Fig. 1c)** or with the hGH tested on the Nalm6 cells, expressing the MuSK3-28 BCR.

### Generation of MuSK-specific human mAbs from circulating B cells

We developed a flow cytometry panel to isolate MuSK-specific B cells (CD3^-^CD14^-^CD19^+^IgD^-^ CD27^+^IgM^-^MuSK^+^) using the fluorescently labeled MuSK antigen **(Supplement Fig. 2)**. We started with 10^7^ PBMCs from each of twelve samples, derived from nine unique MuSK-MG patients **(Supplement Table 1)**, then after B cell enrichment, sorted single MuSK positive B cells. These single B cells (N=672) were individually cultured using a well-established system that induces differentiation to antibody-secreting cells (ASC) such that the supernatant can be screened for antibody specificity [67]. Screening identified two different MuSK autoantibody-expressing cells isolated from patients MuSK MG-1 and MuSK MG-4 **(Fig. 1a; Table 1).** Index sorting showed that both originated from plasmablasts **(Table 1)**, subclass PCRs showed that both expressed IgG4, and they had acquired somatic mutations including those coding for variable region glycosylation sites. Next, mAbs were generated from the BCRs of these two cells and their MuSK binding specificity was tested. These mAbs are referred to as 2E6 and 6E6; both bound MuSK over a broad range of concentrations (ranging from ≈0.03-10μg/mL) in a live cell-based assay **(Fig. 1b)** and this binding was also verified by a clinical radioimmunoassay **(Supplement Table 2)**. Additionally, binding to MuSK expressed on muscle tissue was tested by immunofluorescent staining of murine neuromuscular junctions. Both mAbs bound to MuSK, closely associated with the AChR at the neuromuscular junction **(Supplement Fig. 3)**.

**Fig. 1:**
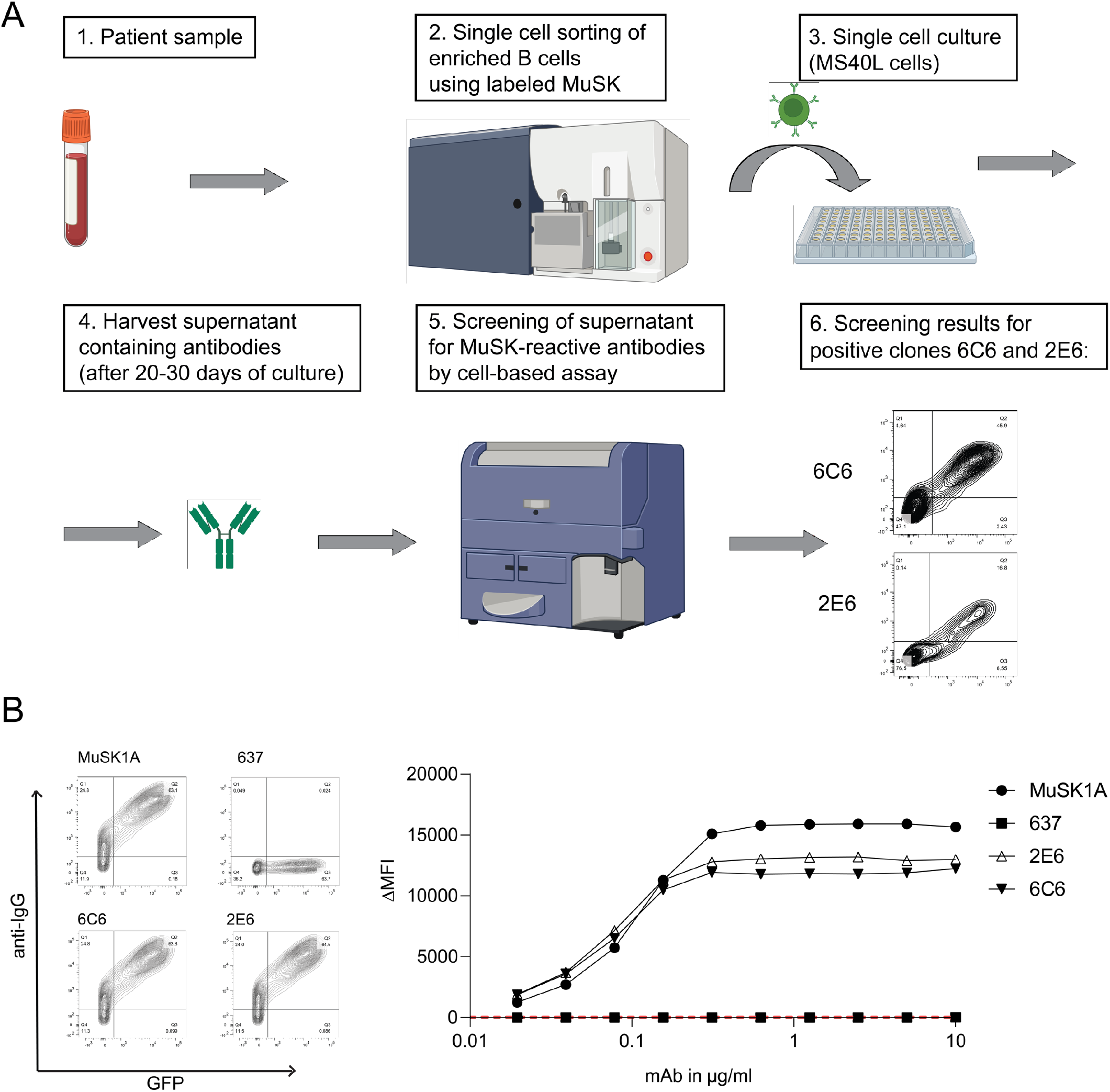
Generation of MuSK-specific mAbs 2E6 and 6C6. **(a)** Illustration of workflow to generate MuSK mAbs. Patient samples were acquired (1) and after B cell enrichment, were sorted (2) into single wells containing a feeder layer for subsequent single cell culture (3). The single cell culture was maintained over a period of 20-30 days, after which the secreted antibodies were harvested (4). A CBA was used to screen each well for MuSK-specific autoantibodies (5). CBA contour plots from this screening show that MuSK-specific IgG are in the supernatant of two culture wells from which the mAbs 2E6 and 6C6 were subsequently derived (6). **(b; left)** Representative MuSK CBA contour plots are shown for the MuSK-specific human mAb MuSK1A, an AChR-specific human mAb 637 used as a negative control, and mAbs 2E6 and 6C6. The x-axis represents GFP fluorescence intensity and, consequently, the fraction of HEK cells transfected with MuSK. The y-axis represents Alexa Fluor 647 fluorescence intensity, which corresponds to the secondary anti–human IgG Fc antibody binding and, consequently, primary antibody binding to MuSK. Hence, transfected cells are located in the right quadrants and cells with MuSK antibody binding in the upper quadrants. The plots show testing with each mAb at a concentration of 1µg/ml. **(b; right)** Binding to MuSK was tested over a series of ten two-fold dilutions of each mAb ranging from 10-0.02 µg/ml. The ΔMFI was calculated by subtracting the signal acquired from non-transfected cells from the signal of transfected cells. Each data point represents the mean value from three independent experiments. Bars or symbols represent means and error bars SDs. Values greater than the mean + 4SD of the negative control mAb at 1.25 µg/ml (indicated by the horizontal dotted line) were considered positive.

**Table 1.**
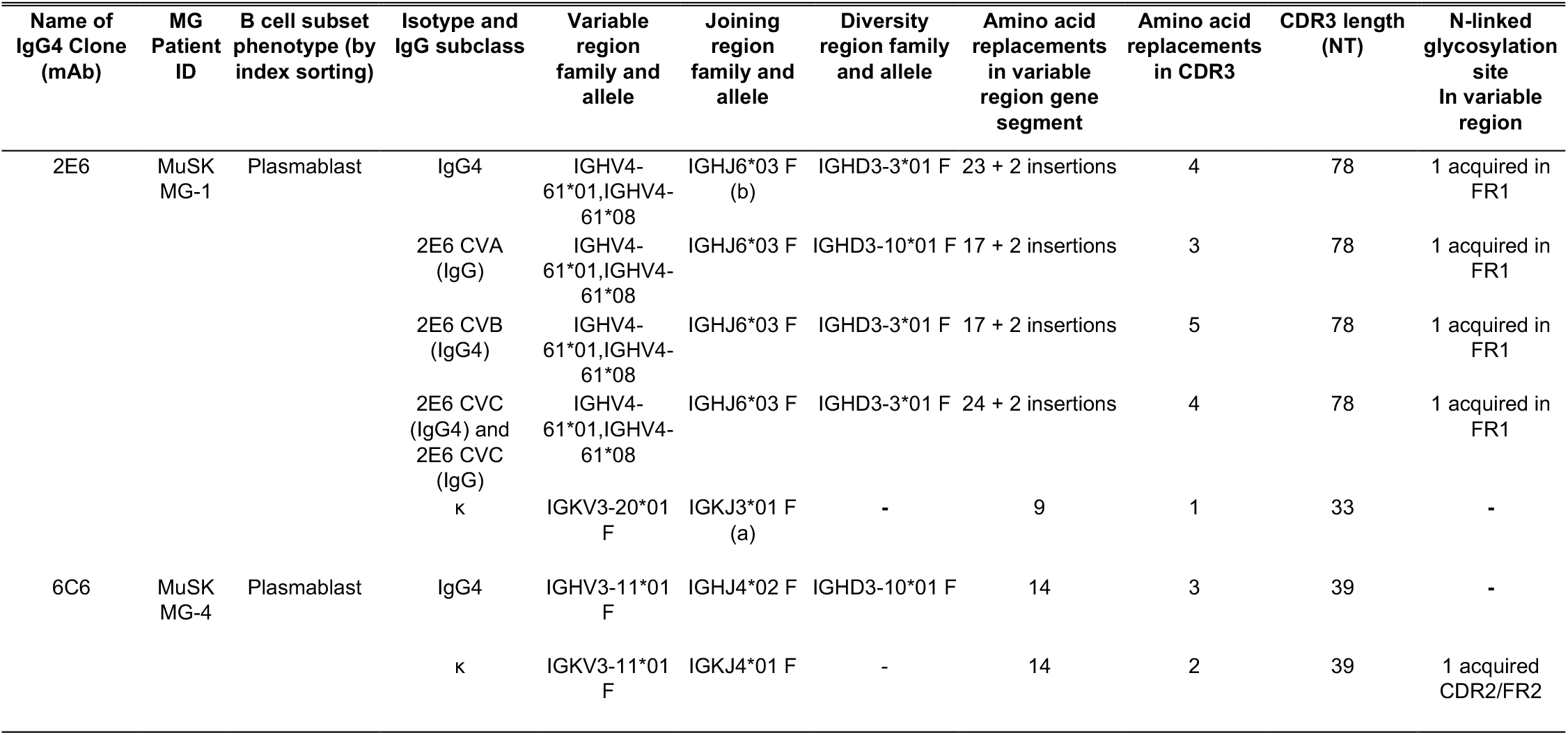
Molecular characteristics of the human MuSK autoantibodies. The molecular characteristics of the human MuSK-specific mAbs 2E6, 6C6 and the clonal variants of 2E6. The replacement mutations in the variable region gene segment were counted from the beginning of framework 1 through the invariable cysteine at position 104. The mutations in the CDR3 were counted between cysteine 104 and the invariable tryptophan (W) or phenylalanine (F) at position 118 in both the heavy chain and the light chain, respectively. No FR4 mutations were observed. N-linked glycosylation motif (N-X-S/T; X is any AA except proline).

### Characterization of MuSK mAbs 2E6 and 6C6

The extracellular domain of MuSK is comprised of three Ig-like domains and a frizzled-like domain **(Fig. 2a)**. Most pathogenic MuSK autoantibodies recognize the Ig1-like domain [29, 30] while a smaller subset bind to the Ig2-like domain [68]. We found that both 2E6 and 6C6 bound to the Ig1-like domain of MuSK and showed no reactivity towards other domains **(Fig 2b, c)**. Valency plays an important role in the pathogenic capacity of MuSK autoantibodies. Monovalent antibodies or Fabs—emulating fab-arm exchanged IgG4—are more potent in disrupting the MuSK-LRP4 interaction which is necessary for the clustering and functionality of the AChR [14, 29, 74]. Therefore, we evaluated the pathogenic capacity of 2E6 and 6C6 as both divalent mAbs and monovalent Fabs by using an established *in vitro* AChR clustering assay [68]. This assay specifically evaluates the capability of autoantibodies to interfere with AChR-clustering, which is dependent upon the MuSK-LRP4 pathway. The number of AChR clusters that formed in response to agrin alone was assigned a value of 100%, and the number that formed in the presence of the mAb or monovalent Fab (tested at equimolar concentrations) was expressed relative to this value. The mAb 2E6 reduced the number of clusters by 53% and mAb 6C6 by 71%. The monovalent Fab of 2E6 reduced the AChR clusters by 61%, while the monovalent Fab of 6C6 reduced the clusters by 96%, similar to the Fab of the positive control MuSK-specific human mAb MuSK1A **(Fig 2d)**.

**Fig. 2:**
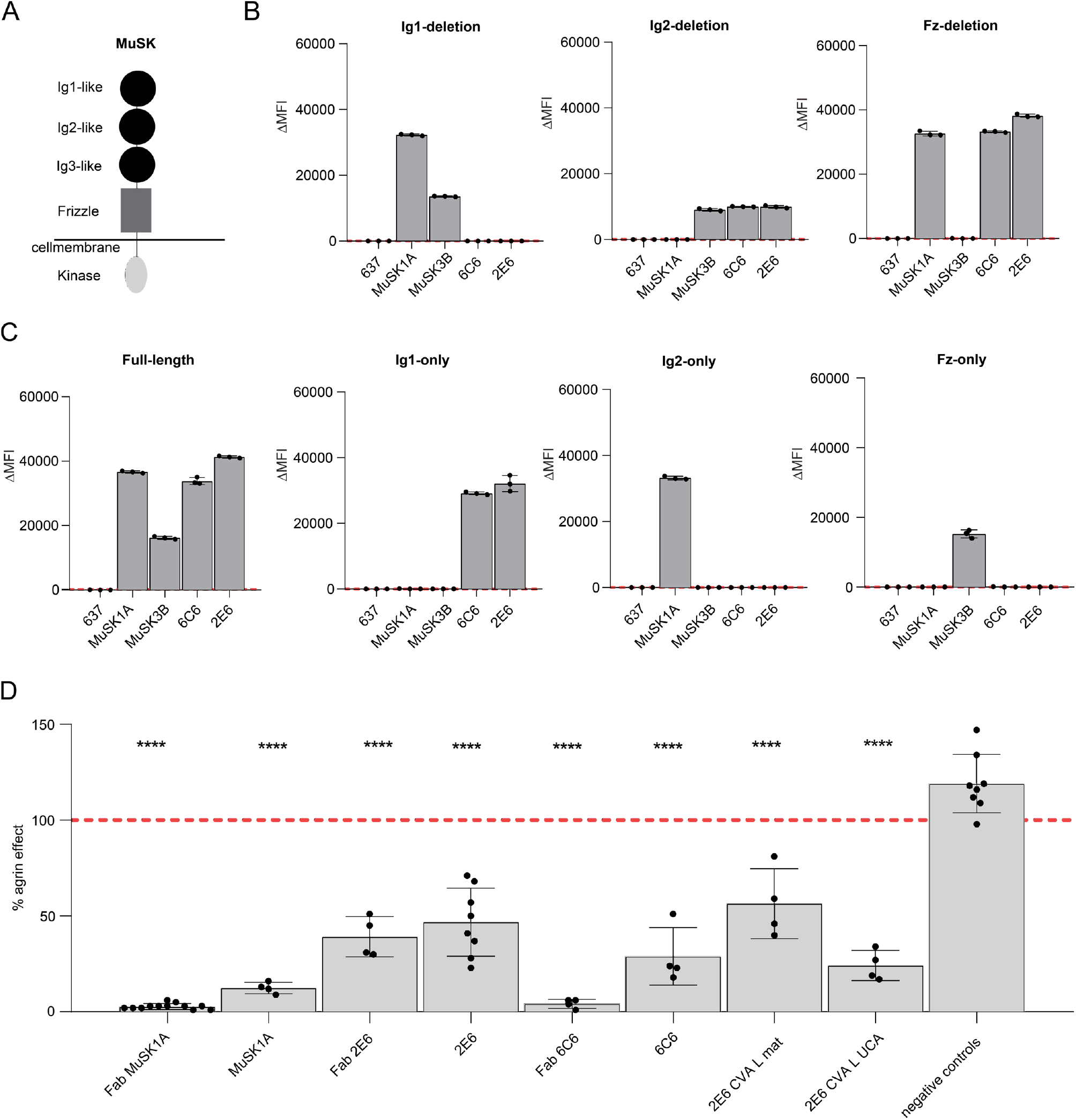
Epitope specificity and pathogenic capacity of MuSK-specific mAbs 2E6 and 6C6. The mAbs 2E6 and 6C6 were tested for domain binding with a CBA expressing MuSK-GFP domain variants. **(a)** Illustration of the full-length MuSK domains. **(b, c)** The ectodomain of MuSK consists of several different Ig-like domains and a frizzled-like domain. Different mutations of the MuSK protein either consisting of a domain deletion or specific domain-only construct were tested for binding by the mAbs. MuSK-specific human mAbs MuSK1A and MuSK3B were used as positive controls (MuSK1A for the Ig2-like domain and MuSK3B for frizzled-like domain) and the AChR-specific human mAb 637 as the negative control. The mAbs were added at a concentration of 10 µg/ml. Results for each mAb are shown. The ΔMFI was calculated by subtracting the signal acquired from non-transfected cells from the signal of transfected cells. Each data point represents a separate replicate within the same experiment, which was measured in triplicate. Bars represent means and error bars SDs. Values greater than the mean + 4SD of the negative control mAb 637, indicated by horizontal dotted lines, were considered positive. **(d)** AChR-clustering assay in C2C12 mouse myotubes demonstrates pathogenic capacity of MuSK mAbs. The presence of agrin in C2C12 myotube cultures leads to dense clustering of AChRs that can be readily visualized with fluorescent α-bungarotoxin and then quantified. Pathogenic MuSK autoantibodies disrupt this clustering. The mAbs 2E6 and 6C6 were tested for their ability to disrupt the AChR clustering. They were tested as divalent mAbs (1μg/mL) and monovalent Fabs (0.3μg/mL). Clonal variant, CVA, of mAb 2E6 was tested with either the mature (mutated) or an unmutated common ancestor (UCA) of the light chain from mAb 2E6, given that the clonal variants were identified with heavy chain-only sequencing. Quantitative measurements of the C2C12 clustering were normalized to the agrin-only effect of each individual experiment. Each data point represents the mean value from 2-8 individual values from a total of 4-10 independent experiments. Bars represent the mean of means and error bars SDs. Multiple comparisons ANOVA (against the pooled results for the three human non-MuSK-specific mAbs), Dunnett’s test; * p<0.05, ** p<0.01, *** p<0.001, **** p<0.0001, only shown when significant.

Unmutated common ancestors (UCA) approximate germline encoded versions of mature antibodies. UCA versions of MuSK mAbs can bind to the cognate self-antigen with high affinity [14]. Therefore, we investigated whether the UCAs of 2E6 and 6C6 can bind to MuSK. We found that the UCA of 6C6 was able to bind to MuSK over a broad range of concentrations (10 - 0.3 μg/ml), while the UCA of 2E6 only showed binding at the highest concentration tested (10 μg/ml) **(Supplement Fig. 4)**. Next, we tested whether the binding of the mAbs and their UCA counterparts is a consequence of polyspecificity **(Supplement Fig. 5)**. We used a well-established approach to test binding to insulin, dsDNA, LPS and a HEp-2 cell lysate [76]. We found that 6C6, the UCA of 6C6 and 2E6 were neither polyreactive nor specific for the HEp-2 lysate, while the UCA of 2E6 was both polyreactive and specific for the HEp-2 lysate **(Supplement Fig. 5a, b)**.

### Phenotype of plasmablasts in MuSK MG at time of relapse after BCDT

The 2E6 and 6C6 mAbs were both isolated from IgG4-expressing plasmablasts during post-BCDT relapse, as were several human MuSK mAbs that we previously produced [65, 68]. Given the increased frequency of plasmablasts in MuSK MG patients during relapse and that these cells can express pathogenic autoantibodies, we sought to explore whether their gene expression profile displayed any unique characteristics, such as expression of pro-survival genes that have been associated with BCDT resistance [9].

To that end, we used single cell gene expression profiling to investigate the phenotype of the plasmablasts at the time of relapse. Single-cell BCR sequencing was performed at the same time to search for clones related to 2E6 and 6C6, and also to explore the MuSK specificity of the IgG4-expressing plasmablast compartment. We used the specific samples from which 2E6 and 6C6 were obtained; MG-1 timepoint 70 months and MG-4 timepoint 25 months respectively (**Supplement Tables 1 and 3**). B cells were sorted to enrich for plasmablasts (CD3^-^CD14^-^ CD19^+^IgD^-^CD27^+^IgM^-^CD38^+^), but with gating that did not entirely exclude other B cell phenotypes. By clustering cells based on gene expression information, we identified 11 distinct clusters, annotated as naïve, memory and plasmablast populations **(Fig. 3 a, b; Supplement Fig. 6)**. We found IgG4 enriched among specific populations in the plasmablast and memory B cell clusters **(Supplement Fig. 6b, 7c)**. Plasmablasts were defined by expression of XBP1, IRF4, PRDM1 and high levels of somatic hypermutations **(Cluster 2,8 and 10; Fig. 3 c, Supplement Fig. 6)**. One plasmablast cluster (number 10) expressed high levels of MKI67, which is associated with proliferation **(Fig. 3 a, b, c)**. We found that CD20 (MS4A1) was expressed at low levels in 25% of the cells within the plasmablast subpopulations, while low level expression of CD19 was found in 25-50% of the population **(Fig. 3 b)**. The expression of high levels of TACI (TNFRSF13B) and BCMA (TNFRSF17) ranged between 25-100% in all three plasmablast subpopulations, while the expression levels of BAFF-R (TNFRSF13C) were low (0-10%) **(Fig. 3 b)**.

**Fig. 3:**
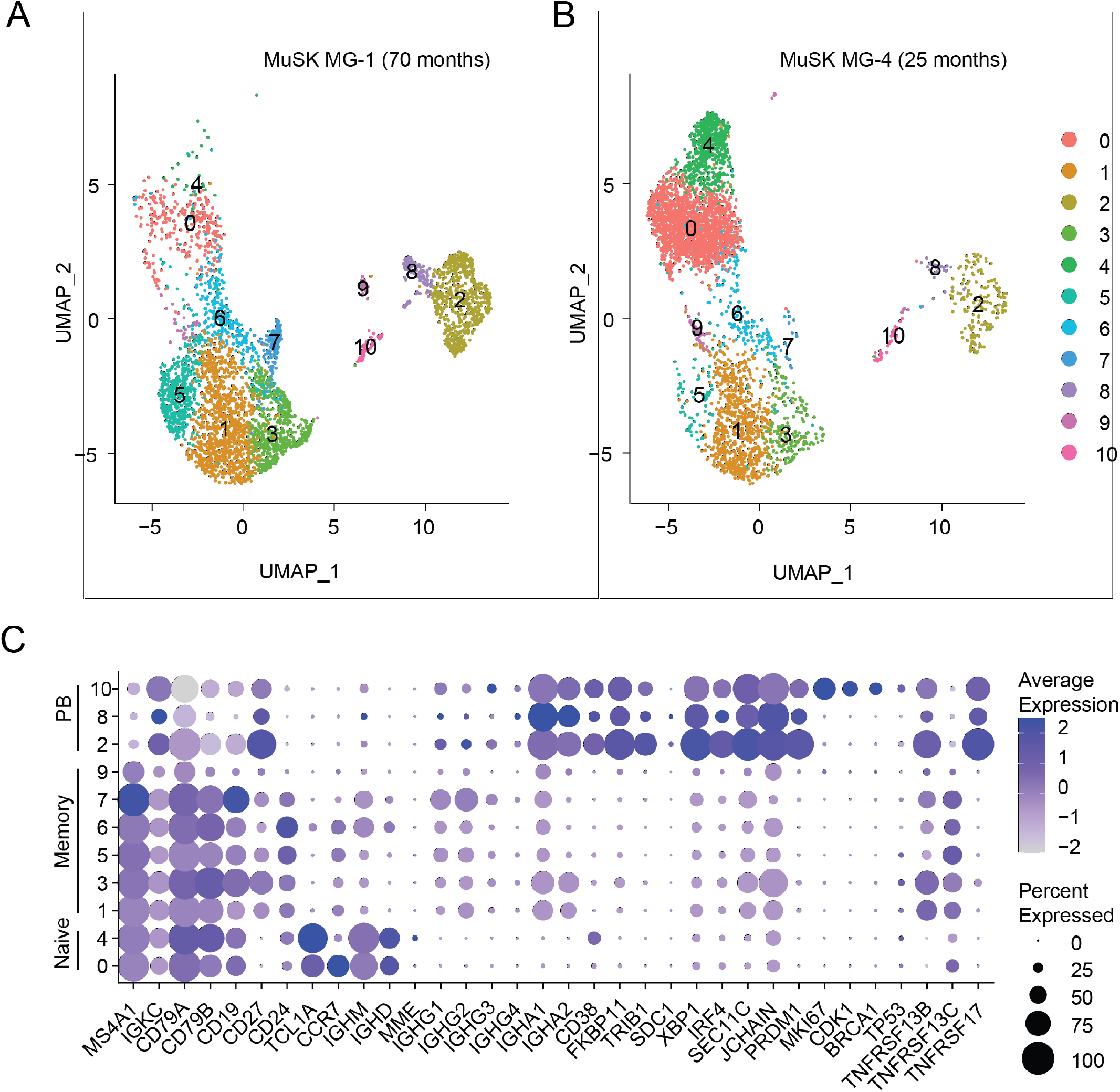
Single cell RNAseq characterization of MuSK MG patient B cells at the time of relapse. Single cell transcriptional profiling was used to characterize B cell samples from patients MG-1 and MG-4 taken at timepoint 70 and 25 months respectively, when MuSK-specific mAbs 2E6 and 6C6 were isolated. Both patients were experiencing a relapse at the time of sample collection. **(a, b)** Uniform Manifold Approximation and Projection (UMAP) of B cells showing 10 populations. Each point represents a single cell. **(c)** Dot plots showing expression of selected B cell marker genes for naive, memory, and plasmablast B cell subsets for the clusters identified in **(a, b)**.

### MuSK-specific B cell clones persist through BCDT and reemerge during relapses

We next investigated whether we could detect historic clonal variants of the pathogenic mAbs 2E6 and 6C6. We had collected serial samples from patient MG-1 (from whom mAb 2E6 was cloned) that made it possible to follow the clinical and treatment course for 79 months, and from patient MG-4 (from whom mAb 6C6 was cloned) for 25 months **(Supplement Table 3)**. We produced BCR repertoire libraries using bulk heavy chain only RNA sequencing and single-cell BCR sequencing to collectively obtain 50,948 unique clones for patient MG-1 and 13,029 clones for patient MG-4 **(Supplement Table 4)**. We identified B cell clones – cells that descend from a common V(D)J rearrangement – by clustering BCR heavy chain sequences by sequence similarity **(Methods, Supplement Fig. 8)**. No historic clones or clonal variants (CV) of mAb 6C6 were found. However, we identified three clonal variants of 2E6, two of which (CVA and CVB) were identified in samples collected prior to that which harbored the plasmablast that produced mAb 2E6, and one (CVC) was identified in the sample that produced mAb 2E6 **(Fig. 4 a; Supplement Table 3)**.

**Fig. 4:**
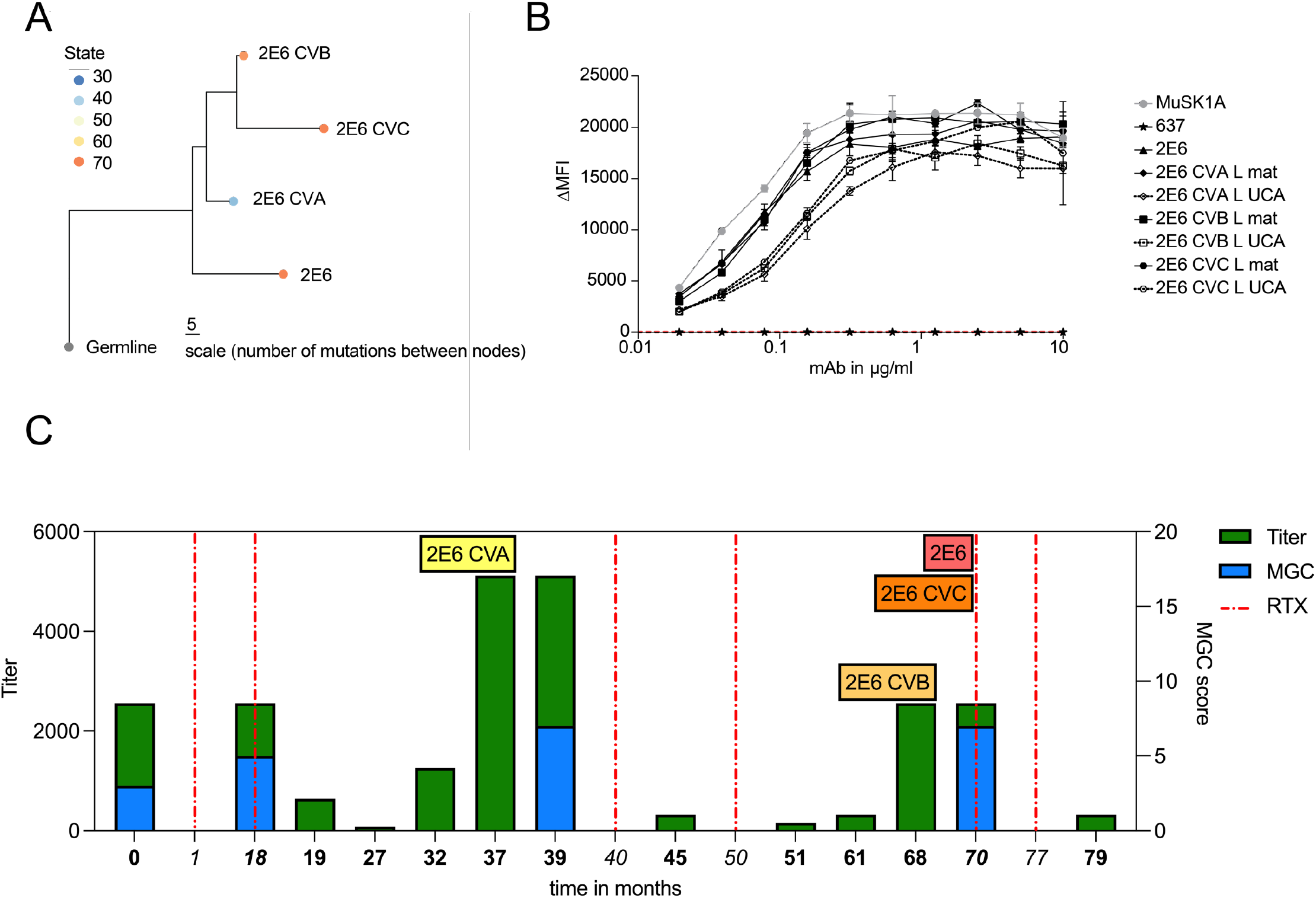
Characteristics of a persistent MuSK-specific B cell clone. Clonal variants of the MuSK-specific mAb 2E6 were identified in longitudinally collected samples from patient MuSK MG-1. **(a)** Clonal lineage containing BCR sequences from bulk RNA sequencing of serial samples collected from patient MuSK MG-1 from whom mAb 2E6 was cloned. A maximum likelihood tree of the 2E6 clone, with clonal variants CVA, CVB, and CVC is shown. Edge lengths represent the expected number of intervening somatic mutations between nodes (see scale bar). Colors correspond to the collection time point (months) at which each clone (sequence) was identified in relation to the first available collection time point. **(b)** Binding to MuSK by mAb 2E6 and clonal variants was tested in a CBA over a series of ten two-fold dilutions of each mAb ranging from 10-0.02 µg/ml. Clonal variants, CVA, CVB and CVC were each tested with either the mature (mutated) or an unmutated common ancestor (UCA) of the light chain from mAb 2E6, given that the clonal variants were identified with heavy chain-only sequencing. The MuSK-specific human mAb MuSK1A was used as the positive control and AChR-specific human mAb 637 as the negative control. The ΔMFI was calculated by subtracting the signal acquired with the non-transfected cells from the signal of transfected cells. Each data point represents the mean value from three independent experiments. Bars or symbols represent means and error bars SDs. Values greater than the mean + 4SD of the negative control mAb at 1.25 µg/ml (indicated by the horizontal dotted line) were considered positive. **(c)** The x-axis depicts the time in months representing the clinical course of patient MuSK MG-1 from whom the mAb 2E6 was isolated. The bold timepoints indicate longitudinal sample collection and clinical assessment; the italicized timepoints indicate BCDT administration. The left y-axis indicates the MuSK autoantibody titer at each timepoint. The autoantibody titer was measured by CBA using 10 two-fold dilutions ranging from 1:20 to 1:10240. The right y-axis shows the MGC score at each timepoint. Dotted vertical lines indicate the timepoints where the patient received rituximab mediated BCDT. Colored boxes indicate the timepoints at which both clonal variants and the original 2E6 mAb were identified. The blue bars show the MGC score, while the green bars show the autoantibody titer.

We next tested whether the other members of the 2E6 clone had pathogenic capacity. Given that bulk-RNA sequencing provided only the heavy chain of each clonal variant, we confirmed the specificity of the variants for MuSK by pairing the heavy chain with either the mature or UCA light chain of 2E6 to approximate the range of light chain sequence variation within the clone. We found that all clonal variants of 2E6 bound to MuSK over a wide range of concentrations when paired with either the mature or UCA light chain of 2E6 (10-0.02 µg/ml) **(Fig. 4 b)**. The mAb of 2E6 originated from a polyspecific germline-encoded UCA antibody that gained specificity towards MuSK over time through affinity maturation **(Supplement Fig. 5)**. Thus, we tested whether the earliest-sampled clonal variant of 2E6 (2E6 CVA) showed pathogenic capacity and specific binding to MuSK. We found that 2E6 CVA showed pathogenic capacity *in vitro* **(Fig. 2 d)**. 2E6 CVA paired with the mature light chain reduced the AChR clusters by 43% and 2E6 CVA paired with the UCA light chain by 76% **(Fig. 2 d)**. Neither recombinant variants of 2E6 CVA (VL mature and VL UCA) were polyreactive, nor did they react to HEp-2 **(Supplement Fig. 5 a, b)**. Both variants bound to MuSK over a wide range of concentrations (10-0.02 µg/ml) **(Fig. 4 b)**. The MuSK MG patient harboring this clone had received rituximab-mediated BCDT 28 months before the patient presented with relapse at the first collection timepoint **(Supplement Table 3)**. The patient received BCDT six additional times over the course of 79 months (6.6 years) and received two cycles of BCDT between the identification of the first clonal variant 2E6 CVA and 2E6 CVB which was 31 months apart **(Fig. 4 c; Supplement Table 3)**. Thus, 2E6 persisted through BCDT and reemerged over time.

MuSK autoantibody titer may be a biomarker for detecting relapse after BCDT-induced remission [69]. In agreement with this finding, we observed that the MuSK autoantibody titer started to increase several months prior to relapse (range: 7-9 months), while the patient was still free of symptoms **(Fig. 4 c; Supplement Table 3)**. The titer remained at the same level during the subsequent relapse and decreased after BCDT **(Fig. 4 c; Supplement Table 3)**. The clonal variants 2E6 CVA and 2E6 CVB were identified at timepoints that preceded relapse by two months **(Fig. 4 c)**. In summary, we found three unique clonal variants of 2E6, which persisted through BCDT. Two of these clonal variants were found during the time at which MuSK autoantibody titer increased and importantly, two months prior to relapse. Together, these results demonstrate the existence of pathogenic B cell clones that survive BCDT and emerge before clinically-detected relapse in MuSK MG.

## Discussion

Most MuSK MG patients respond very well to rituximab-mediated BCDT with a rate of remission of approaching 100% [10, 33]. However, these patients often experience relapse after approximately 1-3.5 years depending on the rituximab treatment regime [7], and a minority of patients do not respond to BCDT therapy [44]. Plasmablast and memory B cell populations are increased at the time of relapse [65, 68] and a subpopulation of these B cells produce MuSK-reactive antibodies [65]. These B cells can, in part, be traced back to clones that existed before BCDT [31]. We previously produced several pathogenic MuSK mAbs [65, 68], but could not identify clones at timepoints other than the time of initial isolation. Therefore, we produced new MuSK mAbs for this current study and used B cell repertoire tracing to search for clonal variants in longitudinally collected clinical samples.

The isolation of the two mAbs, 6C6 and 2E6, was achieved through enriching PBMCs for IgG-expressing memory B cells and plasmablasts that bound to a soluble MuSK antigen. This process required screening of 2.6 x 10^6^ B cells from 12 patient samples; 672 of which were MuSK positive and sorted into plates. Of these 672 sorted cells, two clones were validated and 10 showed high MuSK reactivity, as measured by flow cytometry during the sorting procedure. Although the technical approach is not without limitations, it appears that B cells producing MuSK autoantibodies are exceptionally rare, and that their frequency varies through the course of disease. These findings are consistent with other studies; our own and from other groups who have isolated MuSK mAbs [29, 65, 68]. Furthermore, the number of unique clones in an individual patient appears to be low. In this study we isolated only one clone from each patient and in our other studies we isolated as few as one or two clones from single patients. Although the plasmablast compartment frequency—relative to the B cell population—increases at disease exacerbation, it appears that MuSK-specific plasmablasts make up a very minor population of this expanded compartment. This observation contrasts with findings from studies of acute responses to environmental antigens, where expanded plasmablast clones producing antibodies to COVID or influenza antigens are predominantly antigen-specific and include a diverse clonal repertoire [71, 79]. MuSK autoimmunity may differ in this regard in that only few clones—or even a single clone—could contribute to the production of circulating pathogenic autoantibody, and the expanded plasmablast compartment appears to include many specificities, few of which are MuSK-specific.

The characterization of the MuSK mAbs 2E6 and 6C6 showed that MuSK autoantibodies are heterogenous. Although both MuSK mAbs are not polyreactive, their pathogenic development is quite different. Previously, we had found that the UCA of MuSK mAbs recognize their cognate self-antigen without being polyreactive and that these antibodies develop exceptionally high affinities through the process of affinity maturation necessary to reach their pathogenic potential [14]. These self-reactive antibodies within the naïve B cell repertoire are most likely the consequence of impaired elimination of self-/poly-reactive clones during central and peripheral tolerance checkpoints during B cell development [37, 45, 76]. While the approximated naïve precursor (UCA) of 6C6 does recognize MuSK without being polyreactive—consistent with our previous findings [14]—2E6 develops from a polyreactive precursor gaining specificity for MuSK through affinity maturation. Furthermore, 2E6 and 6C6 differ in terms of the effect of valency on pathogenic capacity. The pathogenic capacity of 6C6 increases as a monovalent Fab, while 2E6 shows similar pathogenic capacity when expressed as a monovalent Fab and divalent mAb. Although it was shown that monovalency increases the pathogenic effect of MuSK antibodies [14, 29, 35, 74], it was also found that an autoantibody requires high affinity towards MuSK to be pathogenic [14]. Therefore, possible explanations for the differing results might be different affinities and binding kinetics of mAbs 2E6 and 6C6, or that heteroligation may be relevant for mAb 2E6.

We demonstrated that the pathogenic 2E6 clone persists through BCDT. The long-term survival of this clone could be the consequence of tissue-based homing, as rituximab is not highly effective in depleting B cells localized in tissues as it is at targeting those in the circulation [2, 36, 42, 55]. Although plasmablasts can express CD20 [31, 52, 65], we found that 25% of plasmablasts in these patients express low levels of CD20 at the time of relapse. Therefore, clone 2E6 might originate from a plasmablast with low expression of CD20 affording it survival through BCDT. Thus, it might be beneficial to use therapies targeting other surface molecules to improve targeting of disease relevant B cell subsets. Recently, new therapies have been developed that deplete B cells, including therapies targeting CD19 [1, 6, 8, 61]. Inebilizumab (anti-CD19) has been approved for the treatment of neuromyelitis optica spectrum disorder (NMOSD) [8], and a clinical trial on the efficacy of Inebilizumab in MG (MINT; ClinicalTrials.gov Identifier: NCT04524273) is currently ongoing. We additionally detected high levels of the receptors TACI and BCMA within the plasmablast subpopulations, but low levels of BAFF-R. TACI and BCMA are part of the BAFF/APRIL-system that regulates the survival of B cells [41, 58]. BAFF-R is the third receptor of the system and the detection of low levels of BAFF-R fits well with previous studies, and is indicative of poor response to treatment with rituximab [3, 31, 59]. The low expression of BAFF-R negatively correlates with BAFF levels [62] and high levels of BAFF are associated with autoimmune diseases, including MG [41]. BAFF levels were shown to be increased in MG [34, 39]. Hence, the BAFF/APRIL axis has been considered as a valuable therapeutic target in the context of autoimmunity and B cell malignancies [38, 66]. Belimumab (anti-BAFF) was already investigated as an add-on therapy in generalized MG patients and showed a subtle positive effect [24]. More recently, chimeric antigen receptor (CAR)-T cells targeting BCMA and/or TACI and anti-BCMA mAbs show promising effects in the treatment of multiple myeloma [38, 54], and autoimmune disease [12], including an ongoing clinical trial in generalized MG (ClinicalTrials.gov Identifier: NCT04146051).

In summary, we generated two new MuSK mAbs that bind to the Ig1-like domain of MuSK and show pathogenic capacity *in vitro*. These autoantibodies were isolated from a sample collected at the time of relapse after BCDT and originated from plasmablasts. The phenotype of the expanded plasmablast population at time of relapse showed variable expression levels of CD20 and CD19, identifying these cells as potential candidates for BCDT, but highlighting that a subpopulation may escape deletion. Clonal variants of a pathogenic, MuSK-specific B cells were identified prior to relapse together with increased MuSK autoantibody levels, both of which may serve as valuable prognostic biomarkers for predicting post-BCDT relapse [69].

## Acknowledgements

We thank Dr. Gianvito Masi for critical reading of the manuscript and Drs. Lesley Devine and Chao Wang in the Yale Flow Cytometry Core for their assistance with cell sorting. The Core is supported in part by an NCI Cancer Center Support Grant # NIH P30 CA016359. We also thank Yale Center for Genome Analysis and Keck Microarray Shared Resource for providing the necessary Pacific Biosciences sequencing services, which is funded in part by the National Institutes of Health instrument grant 1S10OD028669-01.

## Funding support

MLF is supported through a DFG Research fellowship (FI 2471/1-1). KCO is supported by the National Institute of Allergy and Infectious Diseases of the NIH under award numbers R01-AI114780 and R21-AI142198, and through an award provided through the Rare Diseases Clinical Research Consortia of the NIH and MGNet (award number U54-NS115054). SHK is supported by the National Institute of Allergy and Infectious Diseases of the NIH under award number R01AI104739. PMM and MMD were supported by the the Kootstra Talent Fellowship (Fall 2019, Maastricht University) and the ZonMW/NWO Aspasia Program [grant number 015.011.033] and a NIH grant (R21-AI142198). ASP and SO were supported by sponsored research from Cabaletta Bio. The funders had no role in the decision to publish or preparation of the manuscript.

## Competing Interests

KCO has received research support from Alexion, now part of AstraZeneca, and Viela Bio, now part of Horizon Therapeutics, and Cabaletta Bio. KCO is a consultant and equity shareholder of Cabaletta Bio. During the last two years, KCO has served as consultant/advisor for Alexion Pharmaceuticals, now part of AstraZeneca, and for Roche, and he has received speaking fees from Alexion, Roche, Genentech, Viela Bio, now part of Horizon Therapeutics, and UCB. MLF received a SPIN award from Grifols outside the submitted work and is a member of the Alexion-Akademie. KBH receives consulting fees from Prellis Biologics. SHK receives consulting fees from Peraton. ASP has received equity, research support, patent licensing and other payments from Cabaletta Bio; patent licensing payments from Novartis; and consultant fees from Janssen. SO has received patent licensing payments from Cabaletta Bio. The research of PMM and MMD at the School of Mental Health and Neuroscience, Faculty of Health, Medicine and Life Sciences was financially supported by Apellis, Argenx, Genmab, Neurotune and Takeda. PMM is a co-inventor of the following patent: Use of effector-function-deficient antibodies for treatment of auto-immune diseases (Patent number: 9181344). These competing interests played no role in the research design, reference collection, decision to publish, or preparation of the manuscript.

## Notes

### Author Declarations

This study was approved by the Human Investigation Committee at the Yale School of Medicine. Informed written consent was obtained from all patients.

