## Supplemental Data - Figures and Tables for "Reemergence of pathogenic, autoantibody-producing B cell clones in myasthenia gravis following B cell depletion therapy"


| | |
|---|---|
| **Supplement Figure 1** | Validation of a fluorescently labeled MuSK ectodomain reagent to identify MuSK autoantibody-expressing cells. |
| **Supplement Figure 2** | MuSK flow cytometry gating strategy. |
| **Supplement Figure 3** | Staining murine neuromuscular junctions with MuSK mAbs 2E6 and 6C6. |
| **Supplement Figure 4** | Binding properties of unmutated common ancestors from MuSK mAbs 2E6 and 6C6. |
| **Supplement Figure 5** | Polyreactivity and self-reactivity of MuSK mAbs. |
| **Supplement Figure 6** | Somatic mutation frequency and isotype distribution of B cells examined with single cell transcriptional profiling. |
| **Supplement Figure 7** | Distribution of IgG4 expression among B cells clusters identified by single cell transcriptional profiling. |
| **Supplement Figure 8** | Distance-to-nearest plots used to identify the threshold required for assigning clonal members in the BCR sequencing data. |
| **Supplement Table 1** | Study subject clinical, laboratory, and demographic data. |
| **Supplement Table 2** | Radioimmunoassay-based testing of the 2E6 and 6C6 mAbs. |
| **Supplement Table 3** | Characteristics and analysis status of serial samples from patients MuSK MG-1 and MuSK MG-4. |
| **Supplement Table 4** | Counts of reconstructed V(D)J sequences by isotype and clones from sequencing of bulk BCR repertoires and 10x. |

### Supplement Figure 1

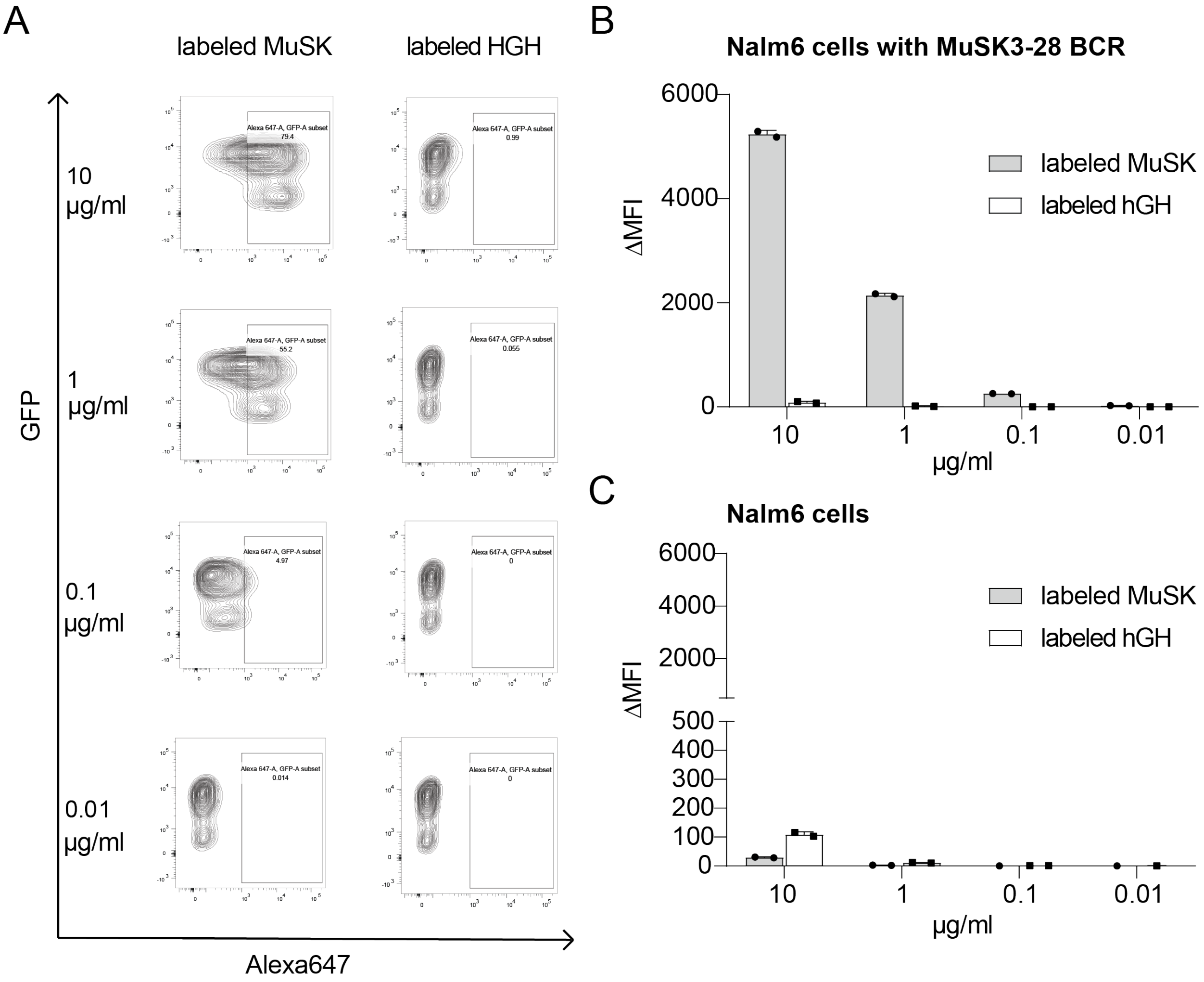


**Supplement Figure 1. Validation of a fluorescently labeled MuSK ectodomain reagent to identify MuSK autoantibody-expressing cells.** A monomeric fluorescently labeled (Alexa Fluor 647) MuSK ectodomain was tested for use in labeling Nalm6-GFP cells, which were engineered to express a B cell receptor (BCR) derived from a MuSK autoantibody (human recombinant mAb MuSK 3-28). Labeled human growth hormone (hGH) was used as negative control. **(a)** Representative flow cytometry contour plots are shown for MuSK and hGH binding to Nalm6-GFP cells expressing a MuSK3-28 BCR. The y-axis represents GFP fluorescence intensity. The x-axis represents Alexa Fluor 647 fluorescence intensity, which corresponds to antigen bound by the BCR. Hence, cells binding to antigen labeled with Alexa Fluor 647 are in the right quadrants. The plots show testing with an antigen (MuSK or hGH) concentration of 10, 1, 0.1 and 0.01 µg/ml. **(b, c)** Binding to antigen was tested with Nalm6-GFP cells expressing the MuSK 3-28 BCR **(b)** or unmodified Nalm6-GFP cells **(c)**. Antigen binding was tested at 10, 1, 0.1 and 0.01 µg/ml. The ΔMFI was calculated by subtracting the signal acquired from testing cells with antigen added from the signal acquired from cells with no antigen added. Each data point represents the mean of value from two independent experiments. Bars or symbols represent means and error bars SDs.

### Supplement Figure 2

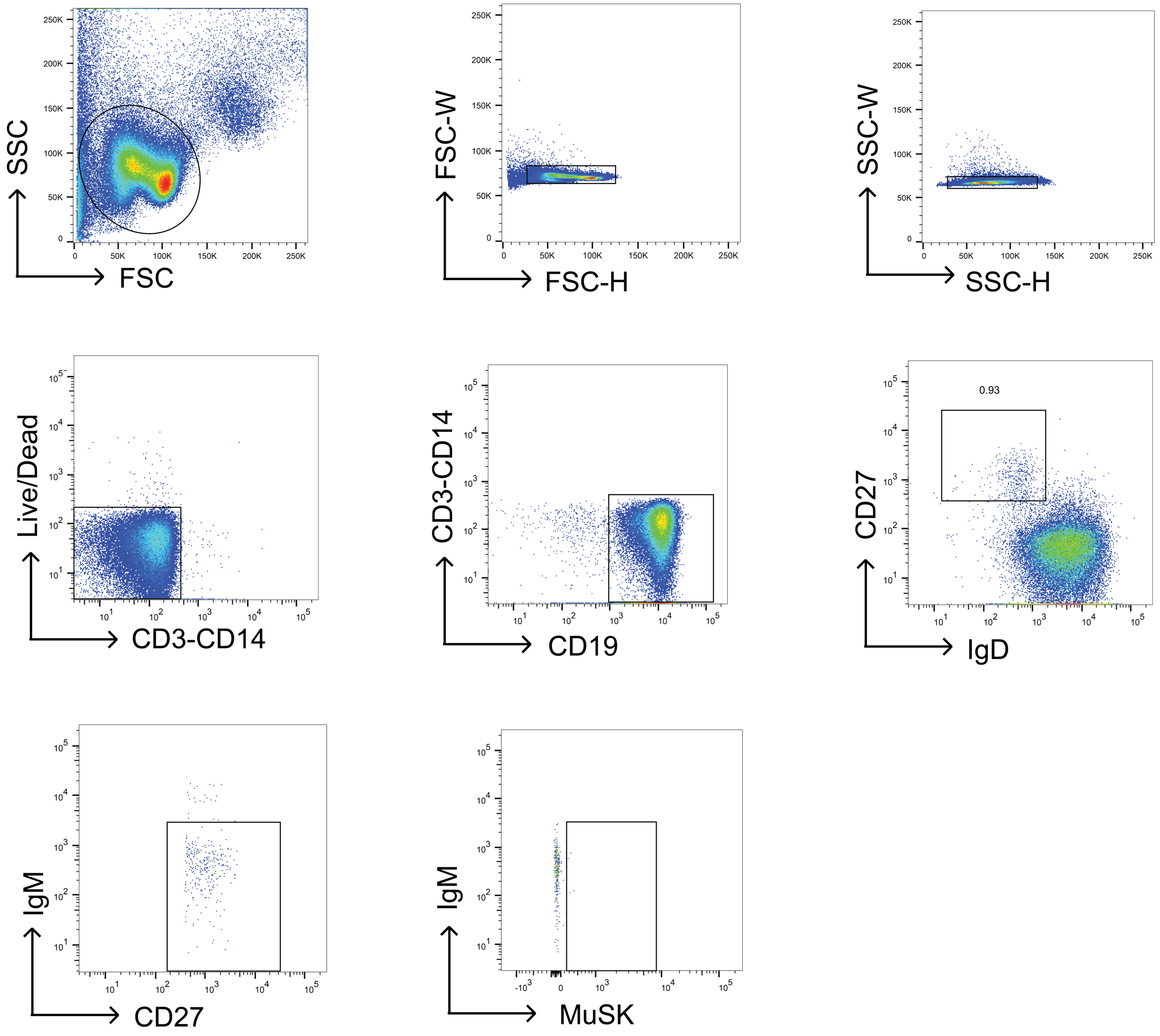


**Supplement Figure 2. Flow cytometry gating strategy for isolation of MuSK-specific B cells.** A representative example of the gating strategy featuring the fluorescently labeled MuSK ectodomain reagent is shown. After B cell enrichment using negative selection beads, single cells were gated using the forward (FSC) and side (SSC) scatter. Dead cells were excluded, then CD3$^{neg}$ CD14$^{neg}$ CD19$^{+}$ IgD$^{neg}$ CD27$^{+}$ IgM$^{neg}$ MuSK-reagent$^{+}$ cells were single cell sorted for subsequent B cell culture and expansion.

**Supplement Figure 3**

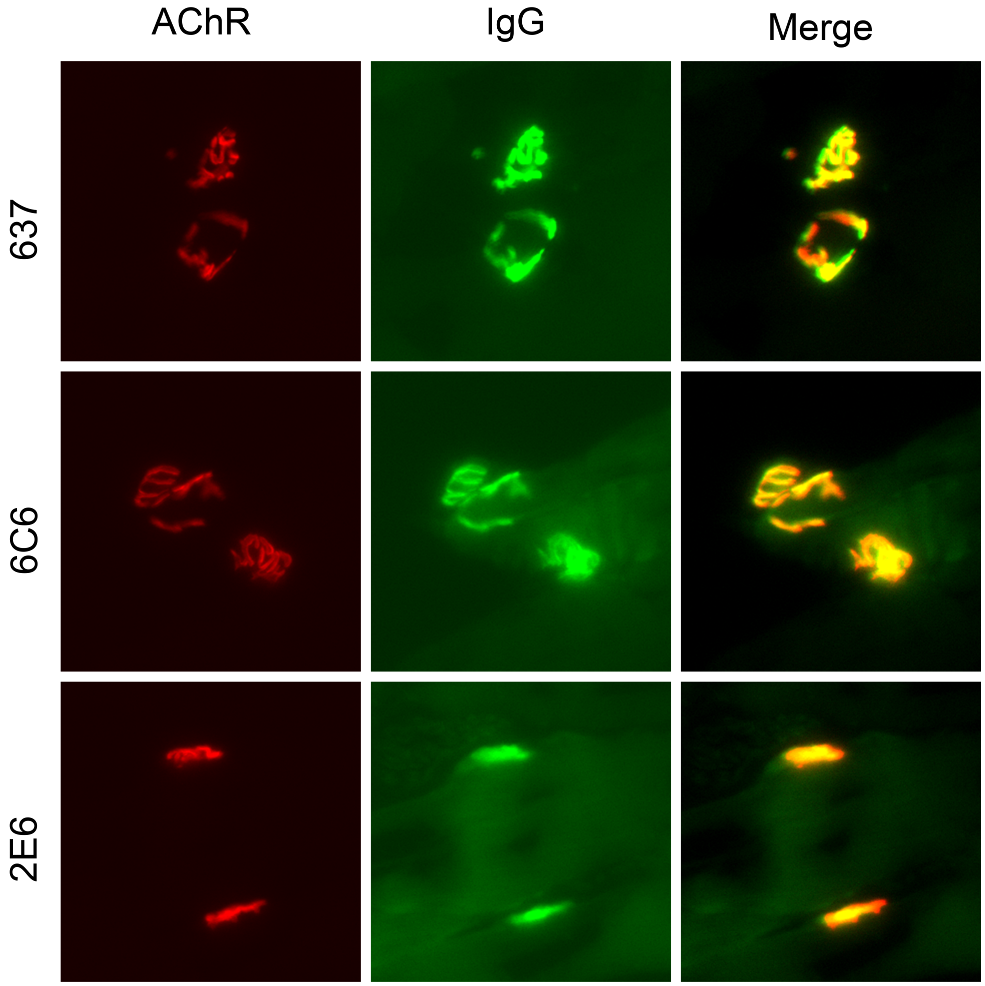


**Supplement Figure 3. Staining murine neuromuscular junctions with MuSK mAbs 2E6 and 6C6.** Immunofluorescent staining of mouse neuromuscular junctions (NMJ). Tibialis anterior muscles were cut longitudinally in cryosections and fixed with PFA. AChRs were stained with Alexa Fluor 648 α-bungarotoxin (shown in red). The mAb 637 was used a positive control, to identify the location of the AChR. Binding of mAbs was detected with goat anti–human IgG Alexa Fluor 488 (IgG, shown in green).

### Supplement Figure 4

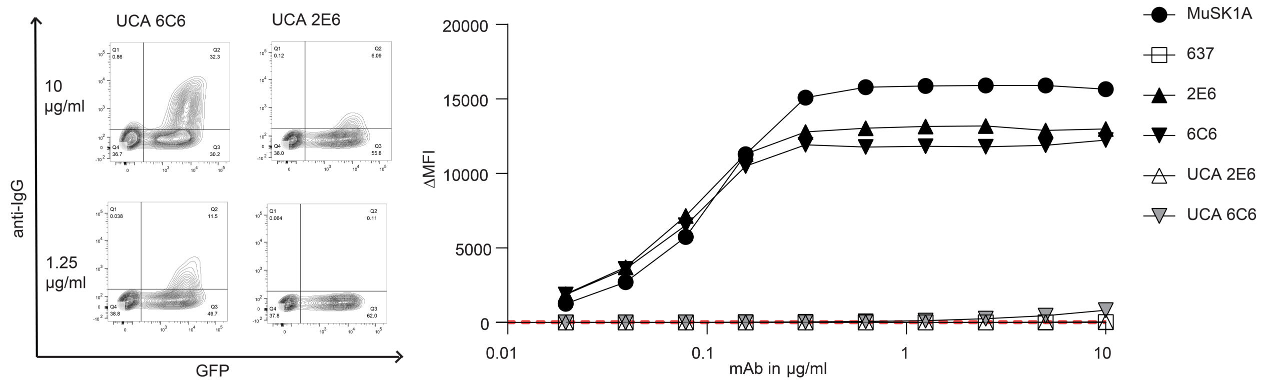


**Supplement Figure 4. Binding properties of unmutated common ancestors from MuSK mAbs 2E6 and 6C6.** Representative cell-based assay (CBA) contour plots are shown **(left)** for the unmutated common ancestors (UCA) of 2E6 and 6C6. The x-axis represents GFP fluorescence intensity and, consequently, the fraction of HEK cells transfected with MuSK. The y-axis represents Alexa Fluor 647 fluorescence intensity, which corresponds to secondary anti–human IgG Fc antibody binding and, consequently, primary antibody binding to MuSK. Hence, transfected cells are located in the right quadrants and cells with MuSK antibody binding in the upper quadrants. The plots show testing with a mAb concentrations of 10 and 1.25 µg/ml. Binding to MuSK was tested over a series of ten two-fold dilutions of each mAb ranging from 10-0.02 µg/ml **(right)**. The MuSK1A mAb was used as the positive control and AChR-specific mAb 637 as the negative control. The ΔMFI was calculated by subtracting the signal acquired by testing non-transfected cells from the signal acquired by testing transfected cells. Each data point represents the mean value from three independent experiments. Symbols represent means and error bars SDs. Values greater than the mean + 4SD of the negative control mAb at 1.25 µg/ml (indicated by the horizontal dotted line) were considered positive.

### Supplement Figure 5

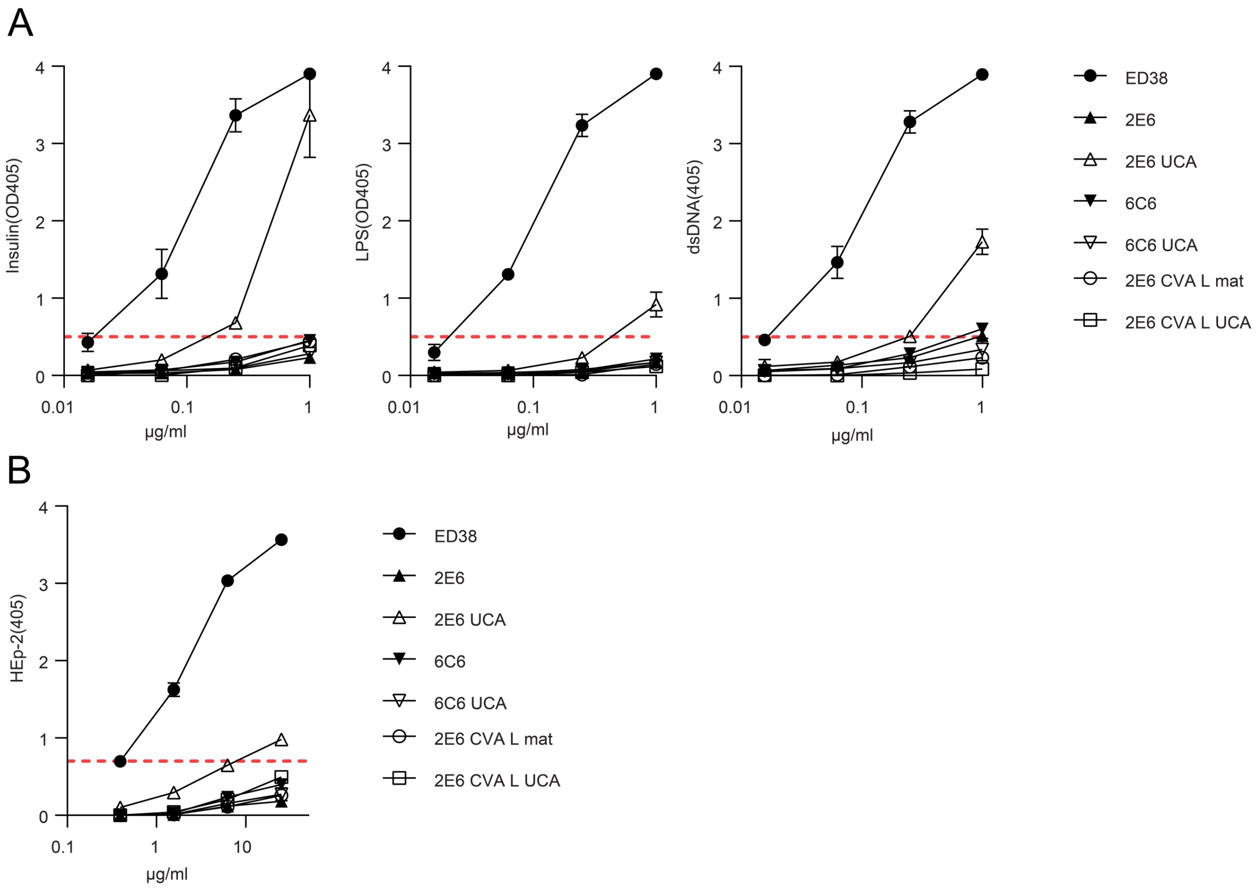


**Supplement Figure 5. Polyreactivity and self-reactivity of MuSK mAbs.** Polyreactivity and HEp-2 reactivity of MuSK specific mAbs 2E6, 6C6 and their corresponding unmutated common ancestors (UCA). **(a)** The reactivity of the mAbs against LPS, dsDNA and insulin was tested by ELISA. Each data point represents the mean value of two independent experiments and the error bars represent SDs. Dotted horizontal red lines mark the positive reactivity cut-off (0.5) at OD405. **(b)** Purified antibodies were tested for autoreactivity on a solid-phase ELISA against human epithelial type 2 (HEp-2) cell lysate. Antibody reactivity to HEp-2 lysate is illustrated by the binding curves. Each data point represents the mean value of two independent experiments and the error bars represent SDs. Dotted horizontal red lines mark the positive reactivity cut-off (0.7) at OD405. ED38, a monoclonal antibody cloned from a VpreB+L+ peripheral B cell, was used as a positive control for both assays.

### Supplement Figure 6

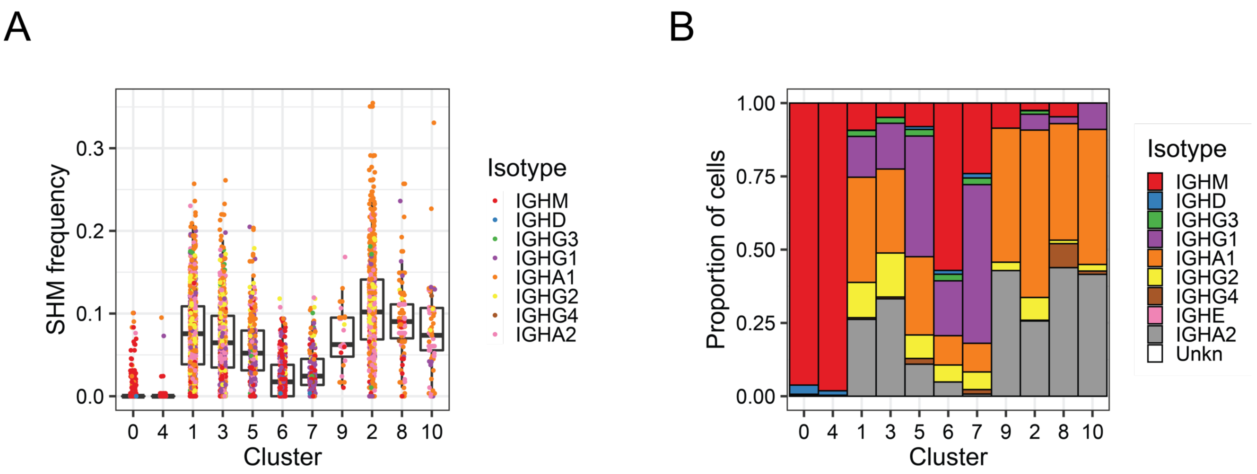


**Supplement Figure 6. Somatic mutation frequency and isotype distribution of B cells examined with single cell transcriptional profiling.** The frequency of somatic mutations **(a)** and isotype frequency **(b)** of the B cell subset clusters shown in Figure 3 a, b.

**Supplement Figure 7**

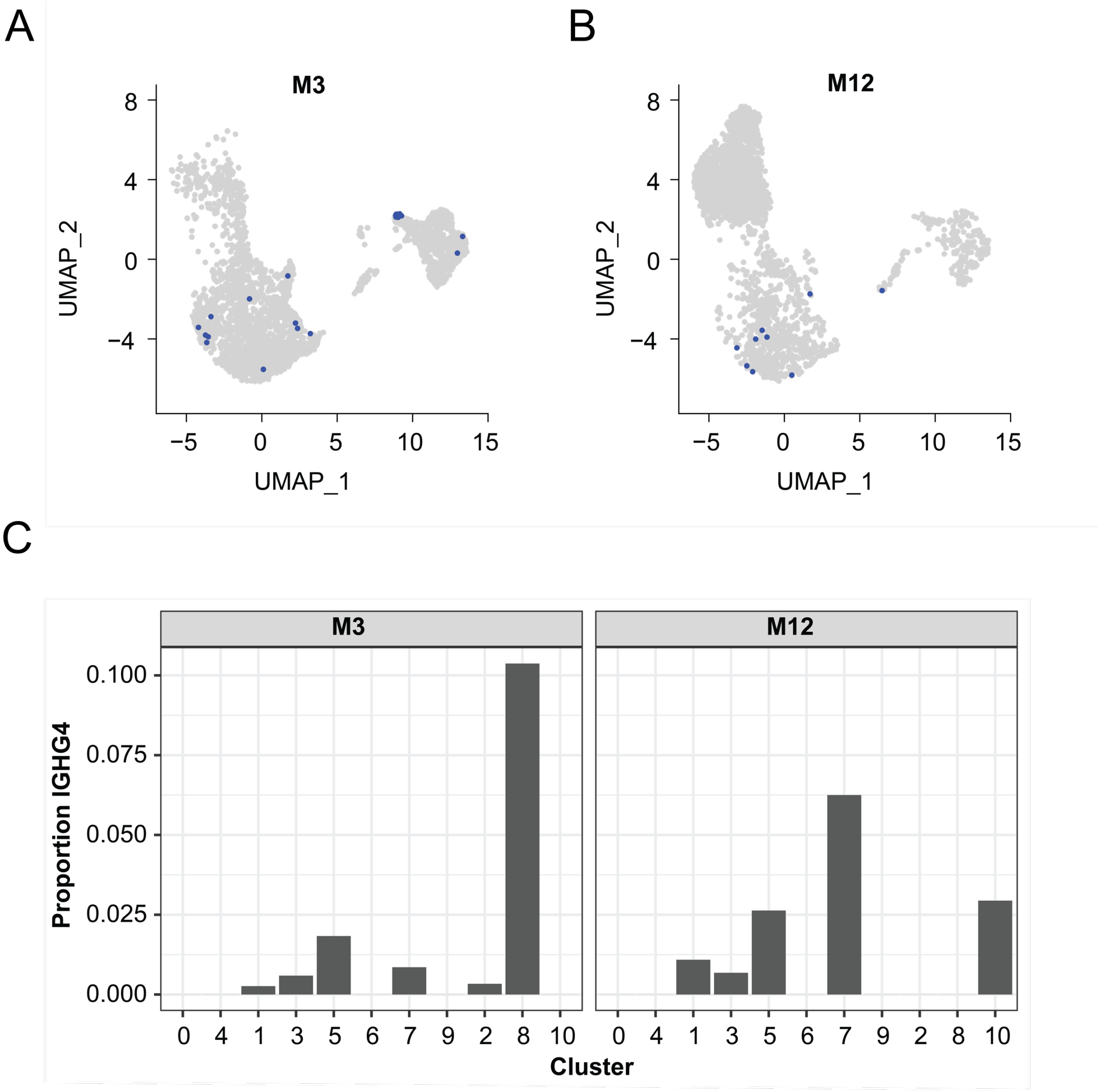


**Supplement Figure 7. Distribution of IgG4 expression among B cells clusters identified by single cell transcriptional profiling. (a, b)** Manifold Approximation and Projection (UMAP) of B cells obtained at the timepoint 2E6 (**a** (=MuSK MG1 (70 months))) and 6C6 (**b** (=MuSK MG-4 (25 months))) were produced. Each point represents a single cell. B cells expressing IgG4 subclass are indicated with a blue dot. **(c)** Proportion of B cells in each cluster shown in Figure 3 a, b with IgG4 constant regions.

### Supplement Figure 8

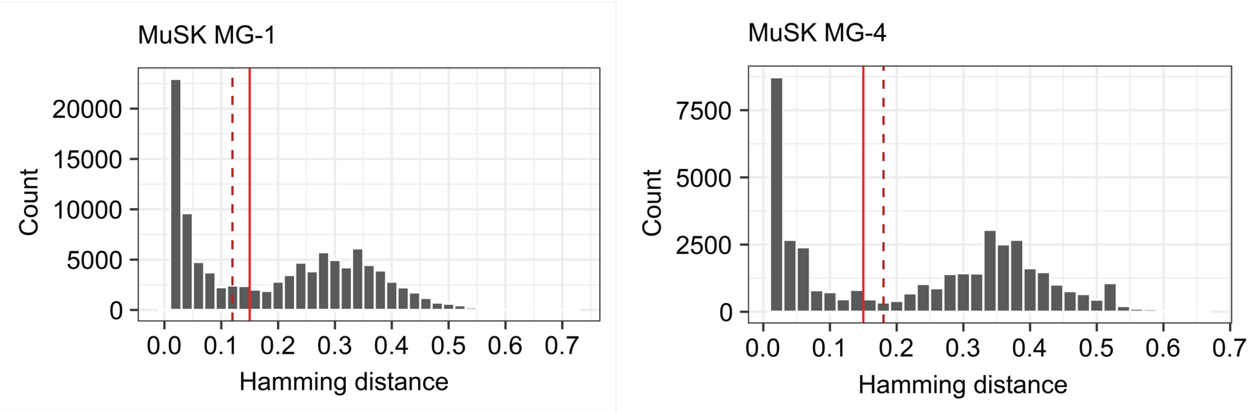


**Supplement Figure 8. Distance-to-nearest plots used to identify the threshold required for assigning clonal members in the BCR sequencing data.** Distance-to-nearest plots used to identify a common threshold to use for hierarchical clustering-based grouping of V(D)J sequences from high throughout sequencing of BCR repertoires. Red dashed lines correspond to the threshold used for assigning clonal clusters. Grey bars represent the distribution of intra-subject distance-to-nearest distances.

| Patient ID | Time (months) since first collection | Sex | Age Range | Diagnosis | Antibody status | MGFA, MGC class at TOC | Time since last rituximab (years) | Other therapy | Serum MuSK antibody titer |
|---|---|---|---|---|---|---|---|---|---|
| MuSK MG-1 | 70 | F | 60-69 | Generalized MuSK MG | MuSK | IIb; 7 | 1.8 | Pred 5 mg/day | 56.0 |
| MuSK MG-2 | 105 | F | 40-49 | Generalized MuSK MG | MuSK | IIb; 2 | 1.5 | None | 12.7 |
| MuSK MG-3 | 47 | F | 50-59 | Generalized MuSK MG | MuSK | IIb; 4 | 0.2 | Pred 10 mg/day | 0.15 |
| MuSK MG-4 | 0 | F | 40-49 | Generalized MuSK MG | MuSK | IIIb; 16 | - | Pred 35 mg/day; IVIg 1g/kg q3wks | 27.3 |
| MuSK MG-4 | 25 | F | 40-49 | Generalized MuSK MG | MuSK | IIIb; 13 | 1.5 | None | 16.5 |
| MuSK MG-5 | 3 | F | 30-39 | Generalized MuSK MG | MuSK | IIB; 7 | - | None | 2.01 |
| MuSK MG-5 | 7 | F | 30-39 | Generalized MuSK MG | MuSK | IIb; 6 | - | PLEX | 8.54 |
| MuSK MG-6 | 0 | M | 60-69 | Generalized MuSK MG | MuSK | IIIa; 15 | - | Pred 60 mg/day; AZA 150 mg/day; Mestinon 60 mg TID; Plex last in Oct 2014 | 1:320* |
| MuSK MG-6 | 16 | M | 70-79 | Generalized MuSK MG | MuSK | IIb; 8 | 0.8 | Pred 10 mg/day; AZA 150 mg/day | 0.03 |
| MuSK MG-7 | 35 | F | 30-39 | Generalized MuSK MG | MuSK | IIb; 4 | 0.9 | MMF 1000 mg BD | 0.89 |
| MuSK MG-8 | 94 | M | 70-79 | Generalized MuSK MG | MuSK | I; 1 | 7.8 | None | 0.0 |
| MuSK MG-9 | 0 | F | 30-39 | Generalized MuSK MG | MuSK | IIb; 5 | - | Pred 40 mg qd | 1.75 |

**Supplement Table 1. Study subject clinical, laboratory, and demographic data.** MuSK MG patients (n=9 unique patients, n=12 different sample collections) from whom samples were used for isolation of MuSK-specific B cells through single cell sorting followed by B cell culturing. Antibody titer was either measured by Athena Diagnostics (unit = fold dilution) or at the Mayo Clinic Laboratory (unit = nmol/L). Samples measured at Athena Diagnostics are indicated by an (*). The reference range for positivity varies according to the measuring facility. For samples measured by Athena Diagnostics the titer range is negative for <1:10, borderline for 1:10 and positive for >1:20. The cut off for negativity for samples measured at Mayo Clinic Laboratory is ≤ 0.02 nmol/L. TOC = time of collection; Pred = Prednisone; PLEX = plasma exchange; AZA = azathioprine; MMF = Mycophenolate mofetil.

| mAb | AChR (CPM) | AChR (ΔCPM) | MuSK (CPM) | MuSK (ΔCPM) | Result |
|---|---|---|---|---|---|
| HC1 | 916 | 153 | 51 | -6 | neg |
| HC2 | 494 | -269 | 62 | 5 | neg |
| HC3 | 878 | 115 | 59 | 2 | neg |
| AChR PC1 | 15908 | 15145 | 81 | 24 | AChR+ |
| AChR PC2 | 18913 | 18150 | 73 | 16 | AChR+ |
| MuSK PC1 | 967 | 204 | 8916 | 8859 | MuSK + |
| MuSK PC2 | 994 | 231 | 8712 | 8655 | MuSK + |
| mAb 2E6 | 667 | -96 | 8268 | 8211 | MuSK + |
| mAb 6C6 | 710 | -53 | 7211 | 7154 | MuSK + |

**Supplement Table 2. Radioimmunoassay-based testing of the 2E6 and 6C6 mAbs.** The radioimmunoassay RIAs were carried out as per manufacturer's instructions. Serum (5 μl) or 5 μl mAb (1 μg/μl) was incubated with 50 μl $^{125}$I-MuSK overnight at 4°C. Carrier was added to mAb tubes followed by 50 μL of anti-human IgG to all tubes for 1 hr. The complex was precipitated by centrifugation, washed, and counted for 1 min on a gamma counter. CPM=counts per minute, ΔCPM=CPM-background, HC=healthy control, PC=positive control.

| Patient ID | Time Point in Months | Diagnosis | Antibody status | MGFA, MGC at TOC | Time since last Rituximab in months | Other therapy | Serum MuSK antibody titer | Sequencing Data available | Origin of Clone or clonal variant |
|---|---|---|---|---|---|---|---|---|---|
| MuSK MG-1 | 0 | Generalized MuSK MG | MuSK | IIb; 3 | 28 | - | 1:2560 | + | - |
| MuSK MG-1 | 18 | Generalized MuSK MG | MuSK | IIb; 5 | 17 | - | 1:2560 | - | - |
| MuSK MG-1 | 19 | Generalized MuSK MG | MuSK | 0; 0 | 1 | - | 1:640 | - | - |
| MuSK MG-1 | 27 | Generalized MuSK MG | MuSK | 0; 0 | 9 | - | 1:80 | - | - |
| MuSK MG-1 | 32 | Generalized MuSK MG | MuSK | 0; 0 | 14 | - | 1:1260 | + | - |
| MuSK MG-1 | 37 | Generalized MuSK MG | MuSK | 0; 0 | 19 | - | 1:5120 | + | 2E6_CVA |
| MuSK MG-1 | 39 | Generalized MuSK MG | MuSK | IIb; 7 | 21 | - | 1:5120 | + | - |
| MuSK MG-1 | 45 | Generalized MuSK MG | MuSK | 0; 0 | 5 | - | 1:320 | - | - |
| MuSK MG-1 | 51 | Generalized MuSK MG | MuSK | 0; 0 | 1 | - | 1:160 | - | - |
| MuSK MG-1 | 61 | Generalized MuSK MG | MuSK | 0; 0 | 11 | - | 1:320 | + | - |
| MuSK MG-1 | 68 | Generalized MuSK MG | MuSK | 0; 0 | 18 | - | 1:2560 | + | 2E6_CVB |
| MuSK MG-1 | 70 | Generalized MuSK MG | MuSK | IIb; 7 | 20 | Pred 5 mg/day | 1:2560 | + | 2E6 2E6_CVC |
| MuSK MG-1 | 79 | Generalized MuSK MG | MuSK | 0; 0 | 2 | - | 1:320 | - | - |
| MuSK MG-4 | 0 | Generalized MuSK MG | MuSK | IIIb; 16 | Pre-rituximab | Pred 35 mg/day; IVIg | 27.3* | + | - |
| MuSK MG-4 | 4 | Generalized MuSK MG | MuSK | IIb; 5 | 2 | Pred 15 mg/day | 25.0* | - | - |
| MuSK MG-4 | 25 | Generalized MuSK MG | MuSK | IIIb; 13 | 18 | - | 16.5* | + | 6C6 |

**Supplement Table 3. Characteristics and analysis status of serial samples from patient MuSK MG-1 and MuSK MG-4.** These longitudinally collected samples were used for investigating whether clones or clonal variants of mAbs 6C6 and 2E6 were present. Antibody titer was either measured by CBA in our laboratory or at the Mayo Clinic Laboratory. Samples of MuSK MG-4 measured at Mayo Clinic Laboratory are indicated by an (*); the unit is nmol/L. The cut off for negativity for samples measured at Mayo Clinic Laboratory is ≤ 0.02 nmol/L. The autoantibody titers of MuSK MG-1 measured by our CBA was performed using 10 two-fold dilutions ranging from 1:20 to 1:10240. TOC = time of collection; Pred = Prednisone

| Patient ID | Time Point in Months | Source | Reads | IGHA | IGHD | IGHE | IGHG | IGHM | Clones |
|---|---|---|---|---|---|---|---|---|---|
| MuSK MG-1 | 0 | Mandel-Brehm et al. 2021 | 7666 | 1514 | 498 | 1 | 529 | 3707 | 5319 |
| MuSK MG-1 | 32 | Mandel-Brehm et al. 2021 | 16117 | 5559 | 1811 | 2 | 1416 | 2697 | 5612 |
| MuSK MG-1 | 37 | Mandel-Brehm et al. 2021 | 15117 | 2102 | 930 | 0 | 699 | 8459 | 10235 |
| MuSK MG-1 | 39 | Mandel-Brehm et al. 2021 | 24834 | 3770 | 2675 | 0 | 1080 | 11571 | 15822 |
| MuSK MG-1 | 61 | PacBio | 9912 | 2878 | 0 | 0 | 1111 | 0 | 2150 |
| MuSK MG-1 | 68 | NEB | 3362 | 1687 | 91 | 0 | 486 | 544 | 1537 |
| MuSK MG-1 | 68 | PacBio | 10659 | 3086 | 0 | 0 | 1106 | 0 | 2736 |
| MuSK MG-1 | 70 | 10x | 3581 | 1575 | 11 | 0 | 711 | 446 | 2218 |
| MuSK MG-1 | 70 | NEB | 5476 | 2697 | 134 | 1 | 694 | 744 | 2247 |
| MuSK MG-1 | 70 | PacBio | 10526 | 2815 | 0 | 0 | 1801 | 0 | 3072 |
| MuSK MG-4 | 0 | NEB | 997 | 379 | 19 | 3 | 311 | 125 | 632 |
| MuSK MG-4 | 0 | PacBio | 17059 | 2955 | 0 | 0 | 1808 | 0 | 3995 |
| MuSK MG-4 | 25 | 10x | 3669 | 499 | 78 | 0 | 329 | 2576 | 3434 |
| MuSK MG-4 | 25 | NEB | 4994 | 1188 | 353 | 1 | 992 | 1448 | 2711 |
| MuSK MG-4 | 25 | PacBio | 15162 | 1924 | 0 | 0 | 1345 | 0 | 2257 |

**Supplement Table 4.** Counts of reconstructed V(D)J sequences by isotype and clones from sequencing of bulk BCR repertoires and single cell transcriptional profiling.
